# Protracted copy number alteration evolution in Barrett’s oesophagus leading to oesophageal cancer

**DOI:** 10.64898/2026.10.16.26365037

**Authors:** Calum Gabbutt, Angel Fernández-Sanromán, Heather Grant, Salvatore Milite, William CH Cross, Nicholas A Wright, Carlo C Maley, Andrea Sottoriva, Giulio Caravagna, Thomas G Paulson, Trevor A Graham

## Abstract

Detection of copy number alterations (CNAs) in Barrett’s oesophagus (BO) predicts oesophageal adenocarcinoma (OAC) risk. When CNAs first occur is unknown. Here we show that CNAs typically form in BO many decades prior to first clinical detection of BO or cancer, and often during childhood. Using a novel molecular clock analysis, we analysed spatiotemporal longitudinal whole genome data from 80 individuals within BO surveillance. Chr 9p loss of heterozygosity (LOH) tended to be the first CNA event typically occurring at age 10-30 years. Genome doubling (GD) also typically originated multiple decades prior to cancer develop. Patients who progressed to OAC had an enrichment of CNAs late in life. CNA rate was more predictive of cancer risk than CNA burden. Analysis of OAC whole genomes confirmed the early-life origins of cancer-associated CNAs. The malignant transformation potential of BO may be set during the first decades of life, enabling early cancer interception.

## Introduction

Barrett’s oesophagus (BO) is the precursor lesion of oesophageal adenocarcinoma (OAC) (Curtius *et al*., 2021). In Barrett’s, the squamous epithelium of the oesophagus is replaced by a columnar epithelium, reminiscent of intestinal epithelium. The presence of BO is associated with a pronounced (>11-fold) increase in relative risk of OAC occurrence (Hvid-Jensen *et al*., 2011) and so in many countries, BO patients are offered enrolment into endoscopic screening programmes that aim to detect early risk factors of OAC (namely dysplasia) (Qumseya *et al*., 2019; Di Pietro *et al*., 2024). However, the absolute annual risk of progression to OAC per patient is low (<0.3%/year (Hvid-Jensen *et al*., 2011)), meaning that for individuals at low risk of cancer, frequent surveillance is “over-treatment” (Esserman *et al*., 2014). Consequently, there is a significant unmet need for effective cancer-risk stratification of BO lesions.

A well-studied and powerful biomarker of OAC risk is the detection of somatic copy number alterations (CNAs) within the BO lesion during endoscopic surveillance. People whose Barrett’s segment has a high burden of CNAs are at significantly elevated risk of OAC (Reid *et al*., 1987, 1992; Li *et al*., 2015; Timmer *et al*., 2016; Hadjinicolaou *et al*., 2020; Killcoyne *et al*., 2020) and particularly if genome doubling (GD) has been detected (Barrett *et al*., 1999; Rabinovitch *et al*., 2001; Nones *et al*., 2014; Stachler *et al*., 2015; Hadjinicolaou *et al*., 2020). The evolutionary dynamics of CNA-bearing clones, in addition to CNA burden, is also prognostic: large clonal expansions are associated with elevated cancer risk (Maley *et al*., 2004), and similarly so is a higher level of within-lesion diversity of CNA-bearing clones (Maley *et al*., 2006; Martinez *et al*., 2016). This is suggestive that the underlying genome instability (rate of new CNA generation) may also be a prognostic factor. However, the CNA rate is challenging to measure directly as it requires comparing the CNA burden between longitudinal samples.

The dynamics of CNAs in BO early in life – before endoscopic surveillance is initiated - remain uncharacterised. Consequently, how early-life CNA dynamics relate to subsequent cancer risk is also unknown. The natural history of BO is enigmatic: the typical age of onset of BO is not known, but it of course must be prior to clinical detection (typically in middle age or later life (Coleman *et al*., 2011) and could be years or even decades prior. Whether or not CNAs begin to accrue prior to BO formation, immediately after BO formation, or only after a protracted period of time post-BO establishment, is therefore unknown. In most healthy normal tissues somatic clonally-expanded CNAs are very rare, and somatic aneuploidy is predominantly detected in neoplastic lesions (Martincorena *et al*., 2015; Martincorena, 2019): oesophagus is no exception (Ross-Innes *et al*., 2015; Martincorena *et al*., 2018). However, recent single cell genome whole sequencing of normal breast from both BRCA-mutant carriers and wild-type individuals detected many isolated aneuploid cells (Williams *et al*., 2024), and analysis of ovarian cancers shows that GD is an ongoing process occurring repeatedly in different cells few of which expand to detectable levels (McPherson *et al*., 2025). BO, as a premalignant lesion where chromosomal instability often evolves, provides a unique opportunity to characterise CNA evolution in a non-cancer setting.

Here, we hypothesised that resolution of the early-life evolutionary dynamics of CNAs in Barrett’s oesophagus would provide new insights in OAC natural history and enable better cancer-risk stratification. To resolve CNA evolutionary history, we constructed a novel computational tool “CARBINE” that infers calendar-year CNAs occurrence times through analysis of a molecular clock signal from single nucleotide variants (SNVs) accrued with CNA regions within clonally expanded tissue. CARBINE was applied to our previously-published deep whole genome sequencing (WGS) data collected from multiple biopsies at two or three endoscopy timepoints from n=80 BO patients, half of whom progressed (P) to OAC during surveillance whilst the other were non-progressors (NP) during surveillance (Paulson *et al*., 2022). CARBINE was also applied to 97 OACs with whole genome sequencing data (Campbell *et al*., 2020). CARBINE revealed that CNAs begin to accrue in the first few decades of life in BO patients, sometimes even in the first two decades of life, and typically preceding clinical detection by multiple decades. Clinically, the rate of CNA accrual in BO (accessible with CARBINE using input data from only a single point in time) provides significant additional value in predicting cancer risk over CNA-burden measurements alone. Analysis of OAC genomes confirmed that these early-life dynamics also occurred in the clonal lineage that led to cancer.

## Results

### Study design

At the time of clinical detection, copy number alteration (CNA)-bearing clones have been in the BO lesions for an unknown period of time (Figure 1A). We aimed to infer the patient’s age when a CNA first occurred, and study how the unobservable early-life CNA evolutionary dynamics related to the risk of OAC occurrence.

**Figure 1:**
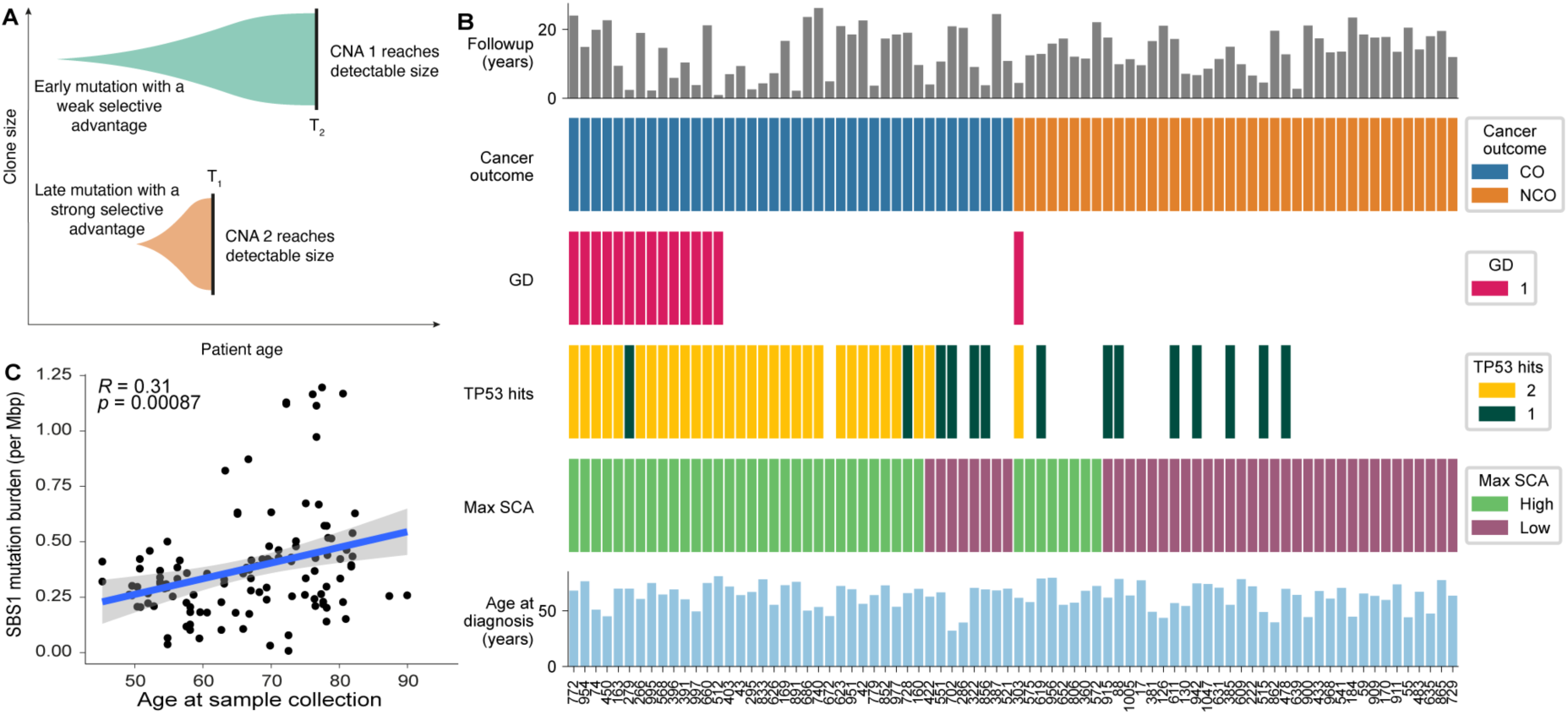
Cohort of 80 Barrett’s oesophagus patients. **A:** Schematic illustrating how the detectability of a genomic alteration (such as a copy number alteration; CNA) in clinical samples is determined by both the time of mutation occurrence and the selective advantage it confers. An early mutation with a weak selective advantage (green) causes a clone to expand slowly and reach detectable size (black bar) only later (T₂), while a later mutation with a strong selective advantage (orange) causes a clone to expand rapidly and reach detectable levels sooner (T₁). **B:** Overview of clinical and genomic features across a patient cohort. Bar plot at the top shows follow-up duration (in years). Below, annotations depict cancer outcome (CO: cancer occurrence; NCO: no cancer occurrence), presence of genome doubling (GD), number of TP53 hits (1 or 2), the maximum somatic copy-number alteration (SCA) burden (split on median SCA, 275.2 Mbp), and age at first endoscopy (years). **C:** Scatter plot showing a positive correlation between patient age at diagnosis and the number of SBS1-associated single-nucleotide variants (SNVs), a signature associated with age-related mutagenesis. The regression line is shown with 95% confidence interval (grey band). A modest but significant correlation is observed (R = 0.31, p = 8.7e-4), consistent with the accumulation of SBS1 mutations over time.

We first assembled CNA and single nucleotide variant (SNV) calls from our previously published whole genome sequencing (WGS) data from n=80 BO patients (table S1). Half of the cohort developed OAC during surveillance (n=40 progressors - P), whereas the other half of the cohort remained cancer free throughout surveillance (n=40 non-progressors - NP), over an equal or longer period of time compared to the progressors (table S2). WGS had been generated from two or more biopsies from two or three endoscopy timepoints per patient (Figure 1B). All WGS profiled biopsies were from non-cancerous BO biopsies. Whole genome library preparation was performed on Barrett’s epithelium isolated from each biopsy, and sequenced to a median depth of 76X [range 40X - 106X] (table S3).

The median burden of CNAs in biopsies from P biopsies (9.4% of the genome, 11.8±1.7 timable CNAs) was significantly higher than in NP biopsies (1.4% of the genome, p=4.7e-11, 1.7±0.2 timable CNAs; Figure 1B). Total SNV burden, and especially the ageing signature (SBS1) C>T mutations, were correlated with patient age at biopsy collection (Figure 1C), confirming previous reports (Alexandrov *et al*., 2015; Martincorena *et al*., 2018) that SBS1 mutations served as a molecular clock in BO/OAC.

### Copy number alteration timing with CARBINE

We constructed a new computational method, “Copy number AlteRation timing with Bayesian Inference and Neutral Evolution” (CARBINE), to calculate calendar-year occurrence of somatic copy number alterations (CNAs) from input whole genome sequencing data from clonally expanded tissue. Like previous approaches (Durinck *et al*., 2011; Greenman *et al*., 2012; Purdom *et al*., 2013; Cross *et al*., 2018; Gerstung *et al*., 2020), CARBINE computes the time of occurrence of a CNA gain by computing the ratio of the number of single nucleotide variants (SNVs) that occurred prior to the gain compared to after the gain. Whether or not an SNV occurred pre or post CNA gain is revealed by SNV variant allele frequency (VAF) because the gain increases the VAF of SNVs on the gained allele (Figure 2A). Compared to previous methods, CARBINE has three mathematical innovations (Figure 2B):

1. Bayesian construction. CARBINE takes prior beliefs about the rate of SNV accrual, clonal expansion rates, and age at which CNAs occur, and performs Bayesian inference upon input whole genome sequencing data to determine posterior distributions (with credible intervals) for these parameters.
2. Hierarchical design. CARBINE shares information on the inferred SNV rate between patients in the cohort to improve the accuracy of parameter inference for cases with limited empirical data.
3. Estimates SNV rate and time since the most recent common ancestor (MRCA) of a sample. The population (clone) of cells within a biopsy were inevitably created through the clonal expansion of a common ancestor cell. This clonal expansion produces a “neutral tail” of low frequency variants with characteristic power-law distribution due to the accrual of passenger mutations within the expanding clone (Williams *et al*., 2016). The exponent of the power-law tail is proportional to the SNV mutation rate per clone size doubling. CARBINE combines the hierarchical design (across patients) and awareness of neutral evolution to learn the SNV rate per calendar year for the input cohort of data. Subsequently, CARBINE is able to infer the time at which the most recent common ancestor of the sampled population emerged.

**Figure 2:**
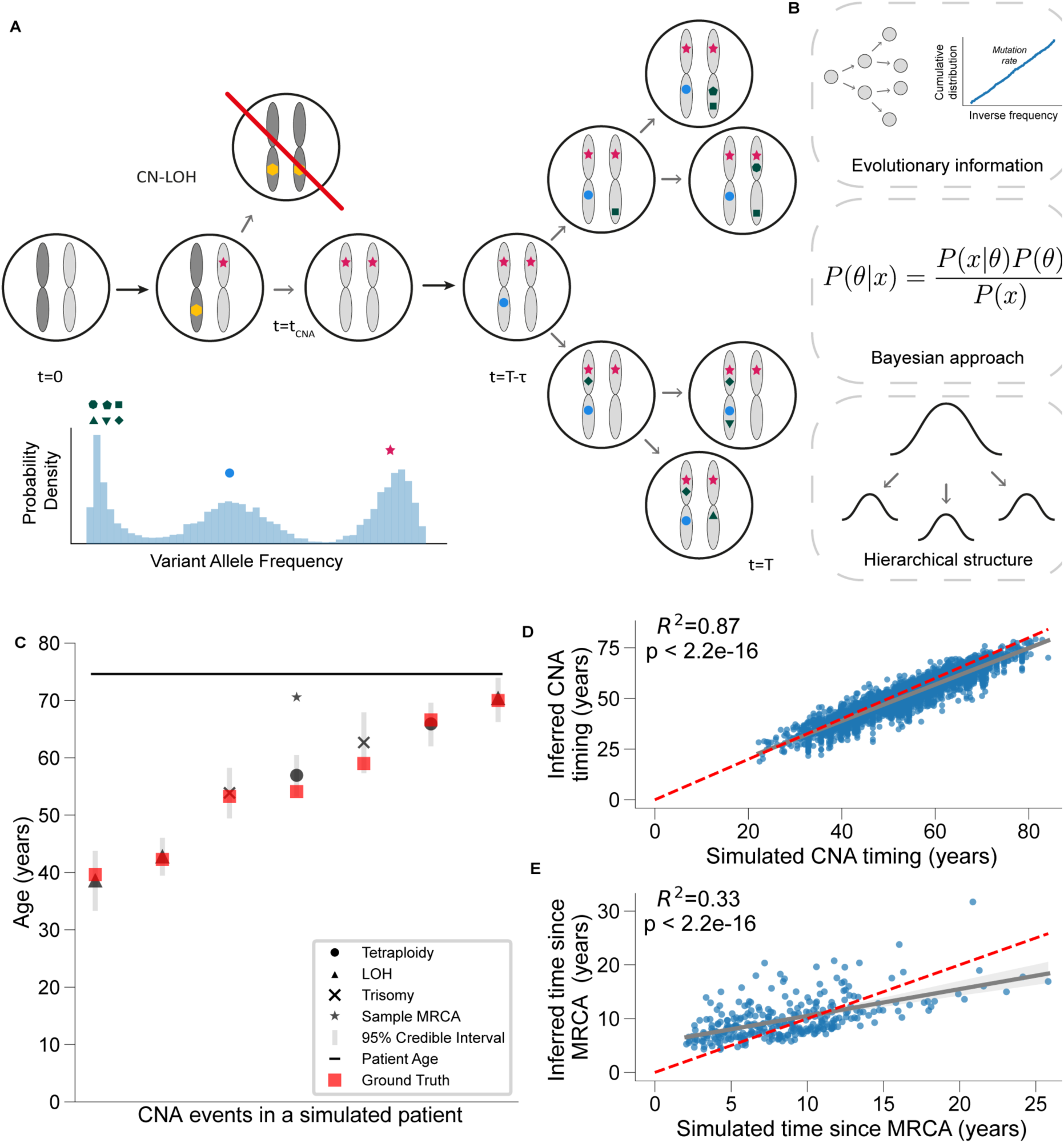
CARBINE incorporates evolutionary information to accurately time copy number gains. **A:** (top) Schematic illustrating how copy-neutral loss of heterozygosity (CN-LOH) and subsequent clonal evolution influence variant allele frequencies (VAFs) and enable inference of the timing of copy number alterations (CNAs). At time t=0, a diploid genome is shown with two parental alleles. At time *t* = *t_CNA_*, a CN-LOH event duplicates the allele containing an early somatic mutation (pink star) and eliminates the homologous region (yellow hexagon), which is followed by the accumulation of additional mutations (blue circle). At time *t* = *T* − *τ*, the most recent common ancestor (MRCA) has emerged, which is followed by clonal expansion with additional mutations arising at decreasing frequency as the population evolves toward the present time point *t* = *T*. (bottom) The histogram below shows the probability density of VAFs for all observed mutations, allowing distinction between amplified clonal mutations (pink star), clonal mutations occurring post-CNA (blue circle), and neutral mutations arising during clonal expansion (green shapes). **B:** Three key methodological advantages of CARBINE. (top) CARBINE leverages patterns of somatic mutation accumulation to inform the sample MRCA timing. (middle) A Bayesian approach to parameter estimation naturally incorporates uncertainty. (bottom) A hierarchical modelling structure allows for the sharing of information across a patient cohort. **C:** An example of running CARBINE on simulated data where the ground truth is known (red). Individual CNA events are presented as separate columns with the posterior mean indicated by a shape (CN-LOH, triangle; trisomy, cross; tetraploidy, circle) and the 95% credible interval represented with a grey bar. The posterior mean of the sample MRCA is represented with a star and the patient age by a horizontal bar. **D-E:** Scatterplot of the simulated CNA time (**D**) and time since the MRCA (**E**) against the inferred value, using the posterior mean as a summary statistic. The least squares fit is overlaid in grey with the 95% confidence interval calculated via bootstrapping. The red dashed line corresponds to perfect agreement between the ground truth and inferred values.

CARBINE uses input data from an entire cohort, in the form of counts of duplicated and non-duplicated SNVs in copy-number gain regions for each sample, along with the inferred effective mutation rates (see Methods). These SNV counts and mutation rates are calculated by applying our subclonal deconvolution tool MOBSTER (Caravagna, Heide, *et al*., 2020; Caravagna, Sanguinetti, *et al*., 2020) to input copy number segmentation data, SNV positions and their variant allele frequencies (VAFs). MOBSTER infers the clonality and multiplicity of SNVs based on their VAF. Explicit handling of the neutral tail and non-neutral subclonal dynamics within MOBSTER prevents incorrect assignment of SNVs that occurred after the sample MRCA (i.e. those in a subclonal peak due to an expanding subclone or in the neutral tail) to single allele copies, and therefore prevents inaccurate CNA timing calculation (Williams *et al*., 2018; Caravagna, Heide, *et al*., 2020).

In its current form, CARBINE only calculated CNA timing for simple gains (gains which occurred in one-step). Regions of complex gains (e.g. where the copy number of a single allele is 3 or higher, which must have been produced by subsequent gain events) were excluded. Across all samples, complex gains accounted for only ∼2.2% of the total genomic regions affected by CNAs, compared to ∼11.1% for simple gains. Code to perform CARBINE timing will be made public via Github prior to publication.

To test the accuracy of CARBINE CNA timing inferences, we constructed a simulation framework where CNAs occurred at random but known times to generate synthetic SNV/CNA data where the ground-truth CNA times were known. Applying CARBINE to these data showed excellent concordance between the inferred data and the ground truth (fig. 2C). The ground truth CNA times and the times since the MRCA were contained within the 95% posterior credible intervals in 100% (4232/4232) of events, with significant tightening of the posterior distributions compared to the prior (supplementary fig. S1). Using the mean of the posterior distribution as a summary statistic, we found significant correlations between the inferred CNA and MRCA times, and the respective ground truths (p<2.2e-16 in both cases, R^2^=0.87 and R^2^=0.33 respectively, fig. 2D-E). Hence CARBINE allows for accurate and precise CNA timing.

Previous tools to infer CNA timing do not explicitly account for the time since the MRCA and the sampling time, nor do they incorporate information from the neutral tail of mutations to help calibrate the timing. To benchmark CARBINE, we ran the widely used CNA timing tool MutationTimeR on our simulated data (Gerstung *et al*., 2020). We found that MutationTimeR systematically overestimated the relative CNA timing and that the correlation between the inferred timing and the ground truth was lower than CARBINE (p<2.2e-16 in both cases, R^2^=0.69 vs R^2^=0.82 respectively, supplementary fig. 2).

### CNAs within Barrett’s oesophagus begin to accrue early in life

We processed the SNV and CNA profiles from our prior study (Paulson *et al*., 2022) using the CNAqc and MOBSTER tools (Caravagna, Heide, *et al*., 2020; Antonello *et al*., 2024), and assigned each SNV to an amplified/non-amplified clonal peak, an expanding subclone or the neutral tail. After merging CNAs of the same type which were located within 1kbp and had a similar ratio of amplified to non-amplified mutations, we had a cohort of 2173 CNAs across 199 samples, covering 71 patients at least once (table S4). As expected, given the known prognostic impact of CNA (Killcoyne *et al*., 2020), progressors had a greater burden of timeable CNAs (1890 vs 283, p<2.2e-16, chi2).

Running CARBINE on these CNAs allowed us to estimate when the CNAs occurred during the lifetime of the patient with high confidence (fig. 3A, median standard deviation on the posterior CNA time was 3.4 years). Remarkably, many CNAs were timed to have occurred early in the lifetime of the patient, with the first timeable CNA per patient often occurring within the first two decades of life (54%, 38/71 patients, fig 3B).

**Figure 3:**
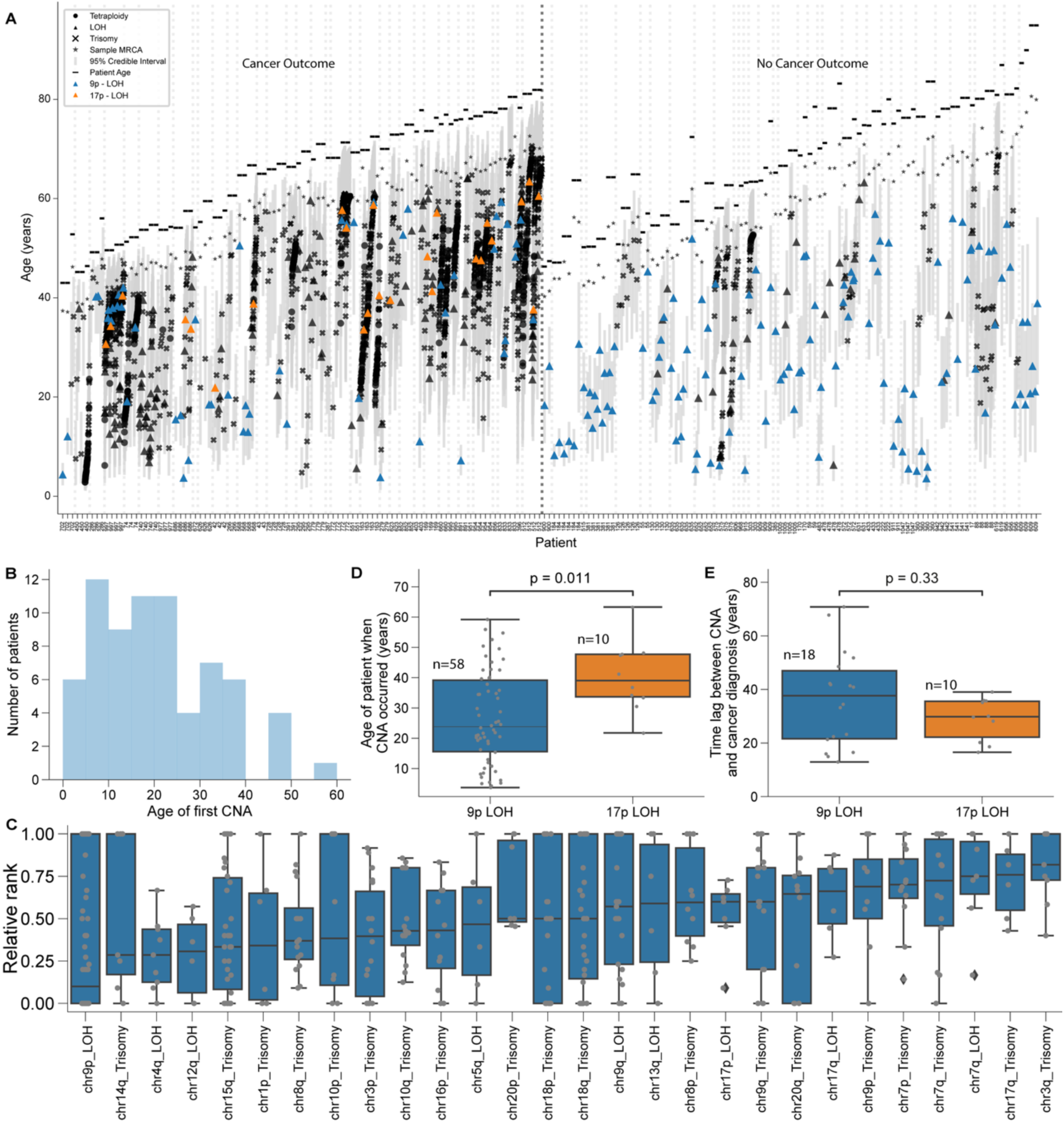
Copy number gains often occur remarkably early in Barrett’s oesophagus. **A:** Copy number alteration (CNA) timings for the 199 samples from 71 BO patients with timable CNAs. Each column represents one sample, and samples are grouped by patient. The posterior mean of individual CNA events are indicated by a shape (CN-LOH, triangle; trisomy, cross; tetraploidy, circle) and the 95% credible interval represented with a grey bar. The posterior mean of the MRCA is represented with a star and the patient age by a horizontal bar. Recurrent 9p LOH and 17p LOH events are annotated in blue or orange respectively. **B:** A histogram showing the age of the patient at the time of the first timable CNA event in any sample. **C:** Box plots showing the relative rank (0 earliest; 1 latest) of recurrent CNAs in non-GD samples, with individual CNAs overlaid as grey dots. Only samples with >1 CNA event are included and only CNAs with >5 occurrences across the cohort are represented. Box-and-whisker plots: centre line, median; box limits, upper and lower quartiles; whiskers, 1.5x interquartile range; points, outlier. **D:** Box plots comparing the age of a patient when 9p LOH and 17p LOH occurred (p=0.011, MW-U test). **E:** Box plots comparing the latency period between either 9p LOH or 17p LOH occurring and the patient progressing to OAC (p=0.33).

One potential complication is that our model depends on the assumption that the mutation rate is constant, which may not be biologically reasonable (Coorens *et al*., 2025). Re-running CARBINE on just clock-like SBS1 and SBS5 SNVs (Alexandrov *et al*., 2015; Paulson *et al*., 2022) demonstrated a high degree of concordance for both the inferred CNA time and the time since the MRCA (p<2.2e-16 for all, supplementary fig. 3). Furthermore, we derived an analytic correction for the inferred CNA time assuming a fixed acceleration in mutation rate following the first CNA event (supplementary note, supplementary figs. 4A-B). Applying this correction factor assuming a 3x mutation rate acceleration yielded qualitatively similar results on our data (supplementary fig. 4C), with the first CNA still occurring within the first 2 decades of life in a quarter of cases (24%, 17/71, supplementary fig. 4D).

Previous work has identified certain recurrent CNAs often present in a characteristic order, with 9p LOH thought to be the initiating event in BO, often occurring as the 2^nd^ hit following *CDKN2A* (9p21.3) mutation (Barrett *et al*., 1996; Galipeau *et al*., 1999). Conversely, 17p LOH often occurs after mutation of TP53 (17p13.1) and is known to increase the risk of progression to OAC (Blount *et al*., 1994; Neshat *et al*., 1994; Reid *et al*., 2001) and GD. In non-GD samples with at least two timeable CNAs, we calculated the relative rank of each CNA and collated across samples (fig 3C). We indeed often observed 9p LOH as the first event, whilst 17p LOH was generally a later event. GATA6 (18q11.2) and ERBB2 (17q12) mutations are also known to be under positive selection in Barrett’s development (Paulson *et al*., 2022), and we observed that 18q trisomy, 17q LOH and 17q trisomy were all observed as late events.

CARBINE allowed access to the real-year timings of CNA acquisition. As expected, 9p LOH occurred significantly earlier than 17p LOH (p=0.011, median 23.8 vs 39.0y, fig. 3D), and occurred anywhere between childhood (earliest event timed at patient age of 4 years) to late-mid-life (60 years old). In the progressor group, there was no statistically significant difference in the time interval between 9p LOH and cancer diagnosis (median 37.7y, range [12.9-70.8y]) compared to the time interval between 17p LOH and cancer diagnosis (median 29.8y, range [16.5-39.1y], p=0.33, fig. 3E), but there was significantly lower variance in the time following 17p LOH than following 9p LOH (p=0.015, Levene test of equal variance). This perhaps suggests that TP53 inactivation (on 17p) acts as a rate limiting step on the pathway towards OAC progression, unlike CDKN2A inactivation (on 9p).

### Samples across time and space validate CNA timings

The multi-sample longitudinal nature of our cohort allowed us to test the concordance of our CNA timings across space and time. We first assessed the clonality of each CNA, classifying each as truncal (present across all samples), shared (present across 2 or more samples, but not every sample) and private (unique to a given sample). As expected, the relative timing of truncal CNAs preceded shared CNAs (p=0.005; fig. 4A), which in turn preceded private CNAs (p=7e-4; fig. 4A).

**Figure 4:**
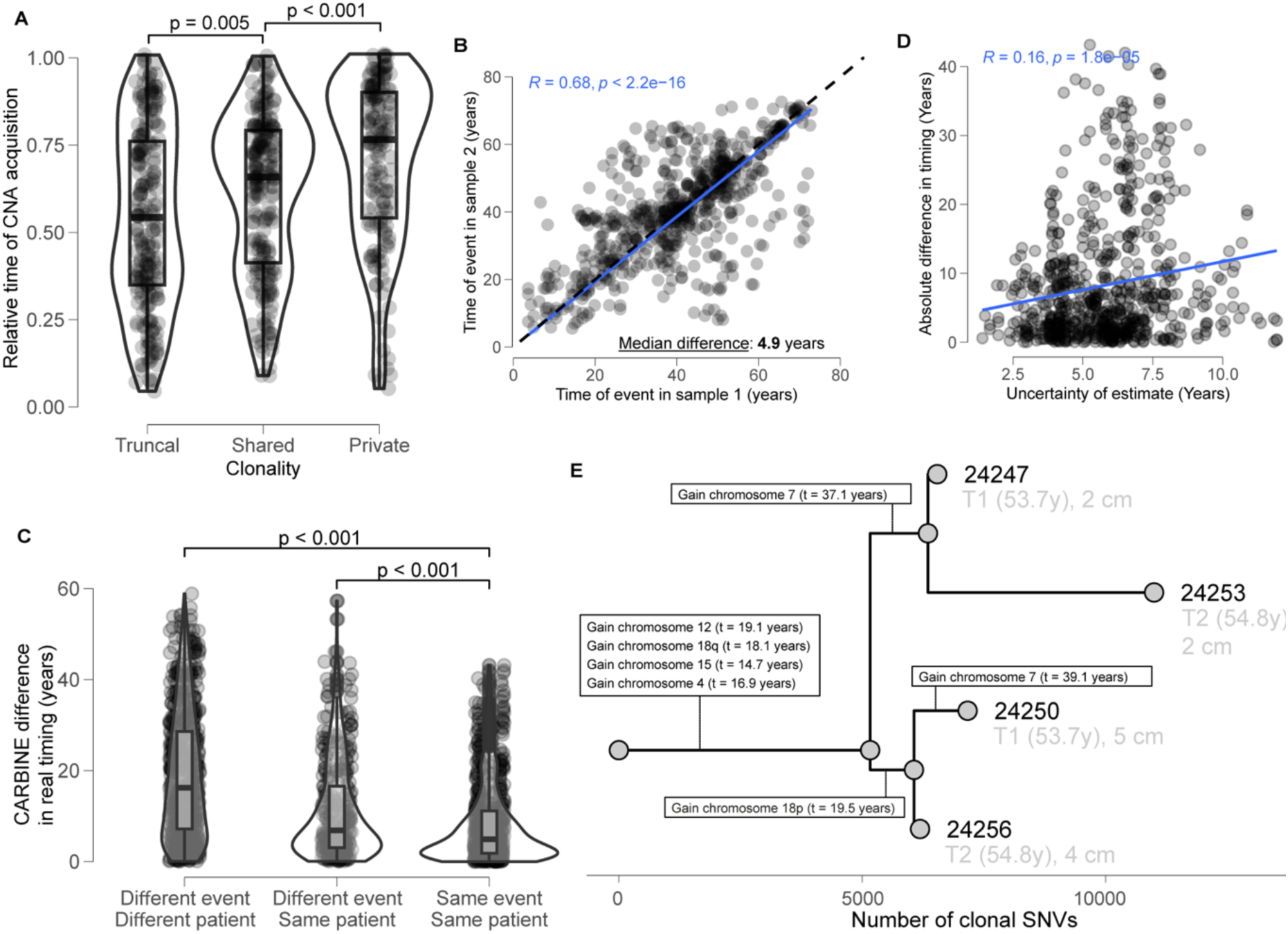
Multi-sample longitudinal data validates CARBINE inference. **A:** Violin plots comparing the relative CNA timing 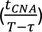 of mutations common to all BO samples from a patient (truncal), CNAs shared across 2 or more but not all samples (shared), and those unique to a single sample (private). Relative timing is significantly earlier for truncal events, supporting the temporal ordering of CARBINE predictions. Violin plots: centre line, median; box limits, upper and lower quartiles; whiskers, 1.5x interquartile range; points, outlier. **B:** Scatter plot showing the timing of matched CNAs across two BO samples from the same patient. Each point represents a CNA timed in both samples. **C:** Violin plots comparing CNA timing differences across various categories. Differences are largest between CNAs from different patients (necessarily not related), smaller between different CNAs from the same patient, and smallest when comparing the same CNA across samples. **D:** Absolute difference in CNA timing estimates between sample pairs from the same patient plotted against the maximum posterior standard deviation of each sample-pairs, a proxy for the uncertainty of those estimates. **E:** Phylogenetic tree constructed from SNVs across four BO samples from a single patient (740). CARBINE estimates of CNA timing (in years) is overlaid onto the tree, showing when specific chromosomal gains occurred relative to the ongoing evolution of the population. The text in grey designates which timepoint the sample was taken (T1 or T2) and the vertical location of the sample in cm with respect to the gastro-oesophageal junction.

Next, we assessed the inferred timings of CNAs that were present (clonal) across more than one biopsy. There was a good correlation between the pairs of CNA timings (p<2.2e-16, R=0.68, fig. 4B), with a median difference of 4.9y. To construct a reasonable null distribution, we calculated the timing distance between every pair of CNAs between samples in different patients and the distance between every pair of different CNAs in the same patient, then compared these to the distance between the same CNA in different samples. The same CNA event in different samples from the same patient had a significantly smaller difference in timing (p<0.001, fig. 4C). We also found a weak but significant correlation between the discordance of shared CNA timings and the uncertainty in the posterior (p<1.8e-5, R=0.16, fig. 4D), suggesting that CARBINE was accurately characterising the inherent uncertainty in the inference.

We compared the consistency of CARBINE timings in the longitudinal samples to the consistency of timings by MutationTimeR run on the same data. MutationTimeR only outputs proportion of evolutionary time for each CNAs, so these were converted to absolute timings by scaling by each patient’s age at sampling. We found that MutationTimeR had a poorer consistency in timing inferences between the same CNA measured in different samples (p<2.2e-16, R=0.57, supplementary fig. 5), with a median difference of 7.6y (vs 4.9y for CARBINE).

The multi-sample cohort also allowed us to place the different CNAs over the phylogeny of the samples we had previously derived from SNVs (from (Paulson *et al*., 2022)) and test whether the CNA timings are consistent with the topology of the tree. In patient 740, the median timing of CNAs occurring on later branches is always later than CNAs occurring on the preceding branches (fig. 4E). In agreement, across the entire cohort, 93 out of 107 (86.9%) private CNAs were estimated to occur later than the median timing of shared CNA acquisition.

In the small number of cases where nominally identical CNAs showed marked timing discordance between samples from the same patient, detailed inspection suggested that these instead multiple independent CNA events at the same locus. In two such patients, discordantly timed 9p LOH events differed in breakpoint structure and in the pattern of shared SNVs across the overlapping 9p region. In patient 568, two samples with early 9p LOH (12.8-12.9y) shared many clonal SNVs, whereas a third sample with a much later timing (50.4y) showed minimal overlap and carried an additional CDKN2A mutation, consistent with a later independent event (supplementary fig. 6). In patient 184, five samples shared an early 9p LOH event (8.2-11.0y), while the outlier sample showed evidence of admixture with a second clone carrying a later 9p LOH (30.6y, supplementary fig. 7). These observations indicate that recurrent 9p LOH can occur multiple times within nested fields of genomically altered oesophageal epithelium.

### The rate of CNA acquisition is variable across samples, patients and time

We calculated the rate of CNA acquisition following the first gain for each sample. Restricting our analysis to non-GD samples, we found that the CNA rates were highly heterogeneous between patients and samples, but that progressor patients had significantly higher CNA acquisition rates (p=1.1e-6, MW-U, fig. 5A), even when accounting for the multiple samples per patient (p=1.3e-3, LMM, supplementary fig. 8A). Interestingly, whilst CNAs in non-progressor samples occurred with approximately equal rate over the lifetime of the patient, there was an enrichment of CNAs occurring just prior to the sample MRCA in progressor samples, leading to estimates of later ages of CNAs in progressors, on average (p=0.012, MW-U, fig. 5B).

**Figure 5:**
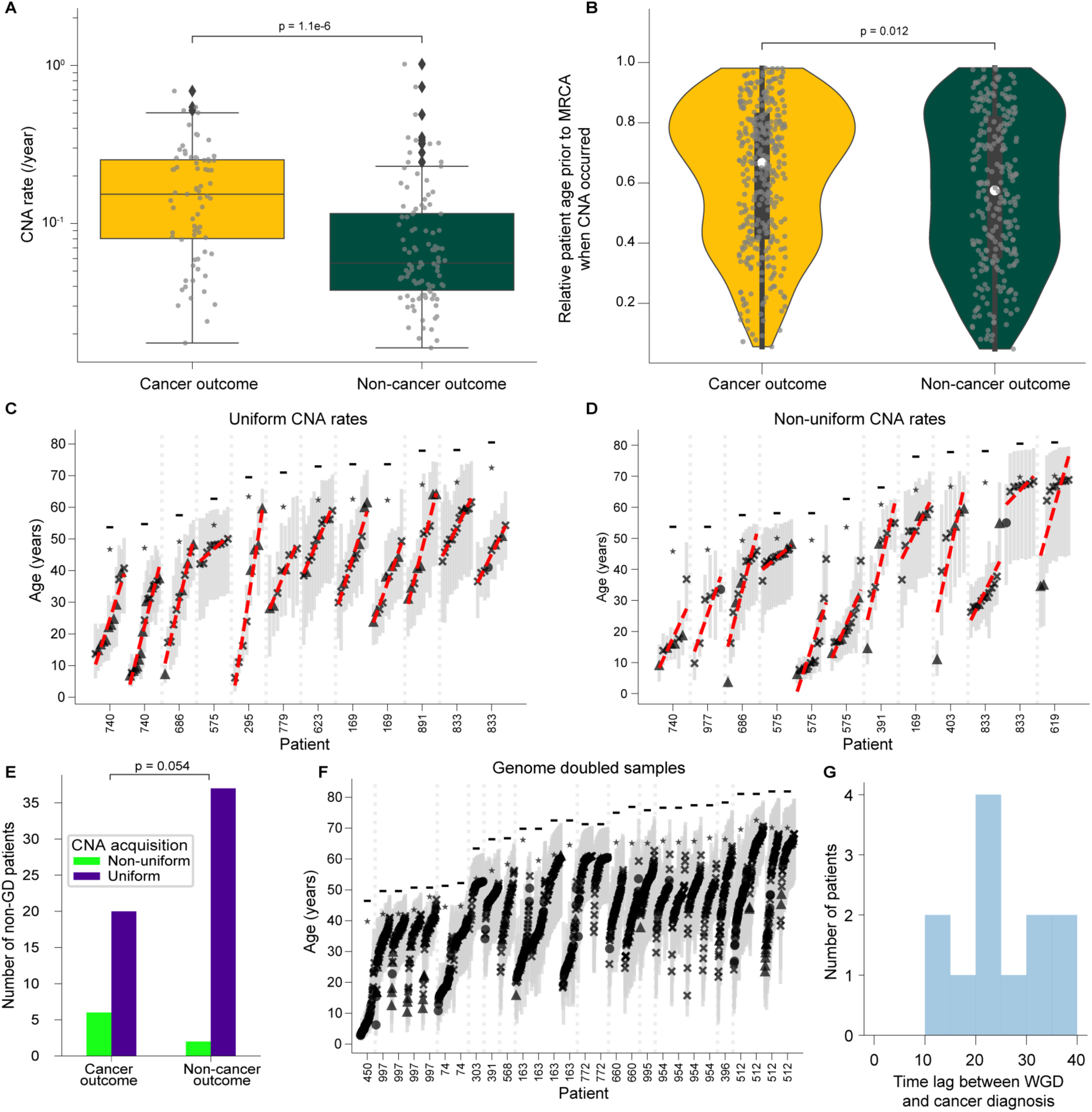
Heterogeneity in CNA acquisition rates. **A:** Box plots showing the number CNA rate (/year) in samples from patients who did/didn’t develop OAC within the study follow-up window. CNA rate was calculated as the number of timable CNA events that occurred divided by the time interval between the first CNA and the age of the patient at the sample MRCA. The CNA rates in individual samples are overlaid with grey dots, and only samples with at least 1 timable CNA are included. Box-and-whisker plot: centre line, median; box limits, upper and lower quartiles; whiskers, 1.5x interquartile range; points, outlier; p value, Mann-Whitney U test. **B:** Violin plots showing the relative CNA timing 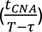 between progressors and non-progressors. Violin plot: centre line, median; box limits, upper and lower quartiles; whiskers, 1.5x interquartile range; points, outlier; p value, Mann-Whitney U test. **C-D:** CNA timings for 12 example samples which were classified as uniform (**C**) and 12 samples that showed evidence of a punctuated accrual of CNAs (**D**). Each column represents one sample, and samples are grouped by patient. The posterior mean of individual CNA events are indicated by a shape (CN-LOH, triangle; trisomy, cross; tetraploidy, circle) and the 95% credible interval represented with a grey bar. The posterior mean of the MRCA is represented with a star and the patient age by a horizontal bar. The best linear fit is overlaid with a red dashed line. **E:** Bar plot showing that the fraction of patients with at least 1 sample with a non-uniform pattern of CNA acquisition vs all uniform samples is higher in progressors than non-progressors (p=0.054, Fisher’s exact test). **F:** CNA timings for genome doubled samples, formatted as in **C-D**. **G:** Histogram of the latency period between GD and diagnosis of OAC. The timing of GD was calculated by taking the median across all 2:1 or 2:2 regions across all GD samples from a patient.

We tested whether the rate of CNA accrual was uniform following the first CNA or followed punctuated dynamics. The distribution of CNA timings was compared to a uniform null hypothesis using either Kolmogorov-Smirnov or *χ*^2^ tests, the results of which were highly correlated (p=9.4e-13, R^2^=0.58, supplementary fig. 8B). 19.6% (12/61) of non-GD samples containing ≥2 CNAs significantly deviated from uniformity (figs. 5C-D). Intriguingly, 23.1% of non-GD patients who progressed to OAC had at least 1 non-uniform sample, compared to only 5.1% of non-progressors (p=0.054, Fisher exact, fig 5E).

Genome doubling is a strong predictor of progression to OAC (Hadjinicolaou *et al*., 2020), often coupled with p53 inactivation. We observed a similar concordance between GD and TP53 mutation - 24/28 samples with GD had one or more TP53 mutations. The TP53 SNV tended to lie in the amplified clonal peak (23/24, supplementary figs. 9-12), confirming that TP53 mutations occurred prior to GD.

We inferred the latency between GD to cancer progression. We assumed that GD only occurred once and therefore estimated the GD time as the median CNA timing of all trisomic and tetraploid segments within the GD samples from a patient (fig. 5F). There was massive heterogeneity in age at GD onset, from 7.2-60.2y, and the time lag between GD and OAC diagnosis was on the order of decades (median 22.3y, fig. 5G). Remarkably, this suggests that GD, rather than occurring just before transformation, instead can be present for very long periods of times, presumably with stalled or very slow clonal expansion, eventually reaching a clinically-detectable size decades later. We note that biopsies represent a tiny fraction of the surface of the Barrett’s tissue and so a small clone would likely be missed by two biopsies at any given timepoint.

### CNA timing in the cancer lineage

To contrast our findings in precancerous tissue with that of cancer itself, we repeated our analysis on 97 deep WGS OAC samples from the Pan-Cancer Analysis of Whole Genomes (PCAWG) consortium (Campbell *et al*., 2020). We ran CARBINE on the 38 samples with sufficiently high purity and data quality (fig. 6A). The first timeable CNA in the cancer genomes often occurred within the first few decades of life (supplementary fig. 13A), as we had observed for CNAs in BO lesion. Unsurprisingly, GD was common within OAC samples (20/38) and we found that GD typically occurred decades prior to OAC diagnosis, with no significant difference between the GD-OAC diagnosis time lag between progressor Barrett’s lesions and OAC samples (p = 0.36, MW-U test, fig. 6B). Within the non-GD OAC samples (18/38), in common with the progressor BO lesions, we found an enrichment of late CNA events (supplementary fig. 13B) and punctuated CNA accrual (10/18 samples, supplementary fig. 13C). Notably, non-GD OAC samples had higher CNA acquisition rates than BO lesions from patients who later progressed to OAC (p=3.0e-7, MW-U, fig. 6C), just as progressor BO lesions had a greater CNA rate than non-progressor lesions (p=1.1e-6, MW-U, fig. 5A). Together, timing CNAs within OAC revealed copy-number dynamics similar to those observed across the whole BO segment prior to transformation.

**Figure 6:**
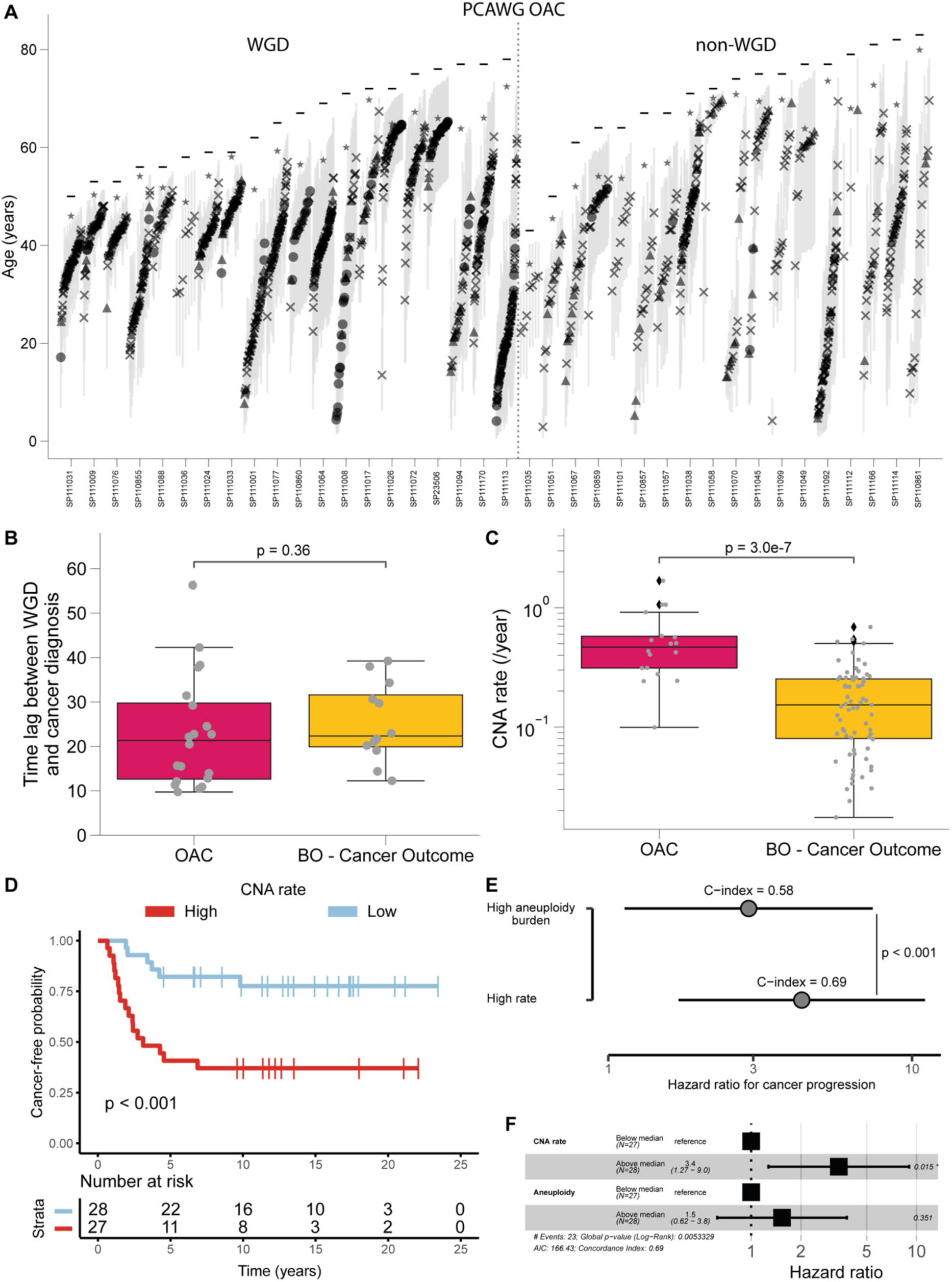
The rate of copy number acquisition is prognostic of progression to OAC. **A:** Copy number alteration (CNA) timings for 38 OAC patient samples from the Pan-Cancer Analysis of Whole Genomes (PCAWG) cohort (Campbell *et al*., 2020) with timable CNAs. Each column represents one patient sample and samples are grouped depending on whether they are genome doubled (left) or not (right). The posterior means of individual CNA events are indicated by a shape (CN-LOH, triangle; trisomy, cross; tetraploidy, circle) and the 95% credible interval represented with a grey bar. The posterior mean of the MRCA is represented with a star and the patient age by a horizontal bar. **B-C:** Bar plots representing the time lag between GD and cancer diagnosis (**B**) and the CNA acquisition rate (**C**) in OAC samples vs samples from BO patients who went on to progress to OAC. Individual patient data points are overlaid (grey) and the difference between groups was assessed via a MW-U test. **D:** Kaplan–Meier analysis of time to progression to oesophageal adenocarcinoma in patients stratified by high (red) or low (blue) CNA rate. CNA acquisition rate was defined as the number of CNA events accumulated from the first detectable CNA to the most recent common ancestor (MRCA). Patients were stratified into high and low groups based on a median split, with the high CNA rate and high aneuploidy burden groups defined as those exceeding the respective cohort medians. P-value was estimated using a log-rank test. **E:** Forest plots illustrating the hazard ratio (HR) for progression to oesophageal adenocarcinoma, estimated via univariate Cox proportional hazards models. Patients were stratified either by aneuploidy burden (top) or by CNA acquisition rate (bottom). Points represent HR point estimates; horizontal lines denote 95% confidence intervals. Concordance indices (C-indices) for each model are displayed above the corresponding HR estimate. The statistical significance of differences in C-index values was evaluated using a U-statistics–based approach for paired survival prediction comparisons. **F:** Forest plot of a multivariate Cox proportional hazards model evaluating the prognostic impact of CNA acquisition rate and aneuploidy burden in the risk of oesophageal adenocarcinoma progression. Hazard ratios and 95% confidence intervals are shown for each covariate, alongside the number of samples in each group and the statistical association with time to oesophageal adenocarcinoma progression.

### CNA dynamics predict clinical outcomes

The burden of somatic CNAs is a well validated prognostic factor for OAC progression in BO (Li *et al*., 2015; Killcoyne *et al*., 2020), as is the presence of GD (Hadjinicolaou *et al*., 2020). Above we found higher CNA accrual rates and an enrichment in CNAs immediately prior to the sample MRCA in progressors (fig. 5A-B), and a further increase in the CNA rate in OAC itself (fig. 6C). Thus, we hypothesised that the dynamics of CNA accrual are more prognostic than sheer burden, due to the additional temporal information that CARBINE revealed. To test this hypothesis, we first randomly selected 1 sample per patient from timepoint T1 and restricted our analysis to only those patients with no history of genome doubling. As expected, the rate of CNA accrual was moderately correlated with aneuploidy burden, defined as the number of megabases subjected to copy-number alterations (p=3.0e-5, R=0.54, supplementary fig. 14A, from (Paulson *et al*., 2022)).

We first confirmed that aneuploidy was itself prognostic in our cohort using both the median and optimal split (p=0.054 and p=0.02 respectively, supplementary figs. 14B-C), the limited sample size notwithstanding. Despite the moderate correlation between patients with a high CNA burden and rate (supplementary fig. 14D), we found that the CNA rate was better for stratifying patient outcomes (p<0.001, fig. 6D), and a direct comparison of the concordance indices of these two factors found that CNA rate was significantly better at predicting progression (p<0.001, fig. 6E). Combining these two factors in a multivariate Cox analysis (with aneuploidy burden and rate binarized into high/low rates) similarly showed that CNA rate remained independently prognostic (HR 3.4 (1.3-9.0), p < 0.015, fig. 6F).

Previous work has identified specific copy number changes as prognostic of the progression from BO to OAC (Paulson *et al*., 2009). To test the interaction between specific CNAs and global genome instability, we tested whether the recurrent CNAs in our cohort were prognostic in these non-GD samples, identifying only 9p LOH, 15q trisomy and 18q trisomy as significant (p=3.0e-4, p=0.0022 and 0.028 respectively, supplementary figs. 15A-C). These CNAs largely overlapped with the CDKN2A, IGF1R and GATA6 genes respectively, which have previously been identified as relevant to BO aetiology (Barrett *et al*., 1996; van Dekken *et al*., 1999; Pavlov *et al*., 2015; Paulson *et al*., 2022). Interestingly, 9p LOH was protective whereas 15q and 18q trisomies conferred a greater risk of progression. However, after including the CNA rate in a multivariate Cox analysis, only 9p LOH remained independently prognostic (p=0.017, supplementary figs. 15D-F). This is due to the correlation between the presence of these specific CNAs and growth rate, where 9p LOH was associated with a lower CNA rate in contrast to a greater growth rate in the 15q and 18q trisomy cases (p=4.9e-4, p=0.0089 and p=0.062 respectively, supplementary figs. 16A-C). Intriguingly, 15q trisomy was often the first timed CNA in patients with many timable CNAs (supplementary fig. 16D), which is suggestive that this event may mark an early branching point into a more chromosomally unstable, high-risk evolutionary path.

## Discussion

Our data reveal extremely protracted evolution of copy number alterations within Barret’s oesophagus lesions. In some patients, the first CNAs are inferred to have occurred in the first years of life. Even though the oesophagus is still growing in childhood (reaching maximum length in late teenage years), it is well-documented that Barrett’s can occur in childhood: Barrett’s is detected in ∼13% of children who have undergone endoscopic assessment following persistent reflux (Dahms and Rothstein, 1984). In fact, one of the hypotheses for the origin of Barrett’s is that some of the columnar cells of the foetal oesophagus do not fully transdifferentiate into squamous cells, and linger near the gastro-oesophageal junction (Wang *et al*., 2011). Given the rarity of CNAs in healthy oesophageal tissue (Martincorena *et al*., 2018), even in later life, our data are therefore strongly suggestive that the development of BO is associated with an increased CNA rate. We cannot tell from our data alone if the increased CNA detection rate in BO is due to an increase in the CNA generation rate, or CNAs experience positive selection (or at least an absence of negative selection) in BO; both mechanisms may operate simultaneously.

Protracted CNA evolution in BO means that there is, in theory, a large window of opportunity for early detection of OAC development risk. It is well established that the detection of CNAs within the Barrett’s lesions is strongly predictive of future cancer risk (Li *et al*., 2015; Killcoyne *et al*., 2020). Here, our data show that these risk-carrying clones often form early in life – increasing the likelihood of future cancer occurrence. Consistent with this finding, our previous analysis of longitudinal genetic data has suggested that clonal expansions are infrequent and slow in BO (Martinez *et al*., 2016). Surprisingly, this is also the case for genome-doubled clones, suggesting that the strong association with cancer risk for GD clones (Barrett *et al*., 1999; Rabinovitch *et al*., 2001; Nones *et al*., 2014; Stachler *et al*., 2015; Hadjinicolaou *et al*., 2020) is only manifest once the clones have grown to sufficient size. Collectively, our data suggest it is plausible that current efforts to implement population screening for Barrett’s (for example through sampling with an oesophageal sponge in a primary care setting (Offman *et al*., 2018) could be augmented with measurement of CNA accrual to simultaneously determine cancer risk. Furthermore, our data show that the evolutionary trajectory of CNAs – namely a high and punctuated CNA occurrence rate – adds substantial prognostic value for early detection of cancer risk. This dynamical information is accessible from single-timepoint data, and so could conceivably be incorporated into contemporary workflows for CNA-based OAC risk prediction (Killcoyne *et al*., 2020).

In addition to genomic biomarkers, DNA methylation biomarkers have also been associated with risk of progression in Barrett’s oesophagus (Wang *et al*., 2009; Agarwal *et al*., 2012; Chettouh *et al*., 2018). The development of enzymatic approaches for joint genetic and epigenetic sequencing raises the intriguing possibility of simultaneously measuring CNA rates and methylome evolution with a single assay (Füllgrabe *et al*., 2023).

Our results indicate that the lineage that ultimately gave rise to invasive OAC exhibits CNA accrual dynamics that closely mirror those of the surrounding BO field, rather than cancer evolution being an “outlier” trajectory. Our previous work has shown that a more genetically diverse BO field (Maley *et al*., 2006; Martinez *et al*., 2016) increases the risk of progression. The shared CNA dynamics between progressor BO and OAC therefore likely reflect that Barrett’s cells in patients who subsequently progress to cancer are inherently prone to acquire CNAs (at a rate higher than in non-progressors) and subsequently develop cancer. This could be because progressors are prone to chromosomal instability (i.e. be due a mutational process) or because CNAs are more strongly selected in progressor lesions (i.e. be due to a selective effect). Future multi-omic profiling and functional studies are required to evaluate these two hypotheses.

Calendar age CNA timing was achieved with a new method “CARBINE” that has three methodological advantages over previous approaches: (a) a Bayesian construction which permits direct handling of uncertainty in timing estimates, (b) a hierarchical design, which enables sharing of information across a cohort of patients to improve inference accuracy in individual patients, and (c) incorporation of evolutionary theory to extract additional information from the data (mutation rate and time elapsed since the common ancestor of a biopsy) and which reduces error in CNA timing inference. The assumptions of CARBINE are general, meaning that the method can be used in any somatic evolution setting featuring clonal expansion.

In summary, calendar-year timing of copy number alterations in Barrett’s oesophagus reveals extremely protracted evolution occurring over decades prior to cancer occurrence, and CNA mutation rate predicts progression to OAC better than CNA burden. This provides a strong rationale for early determination and genomic evaluation of cancer risk in the premalignant disease.

## Methods

### Ethics approval and consent

All research participants contributing clinical data and biospecimens to this study provided written informed consent, subject to oversight by the Fred Hutchinson Cancer Research Center IRB Committee D (Reg ID 5619). All genetic and demographic data used were publicly available prior to the start of the study (Alexandrov *et al*., 2020; Paulson *et al*., 2022).

### Assembly of deep whole genome sequencing (WGS) data

In this study, we utilized previously processed genomic data from our earlier publication (Paulson *et al*., 2022). Briefly, that study included whole-genome sequencing (WGS) of 427 biopsy samples collected from 80 individuals, 40 of whom progressed onto oesophageal adenocarcinoma (cancer outcome; CO) and 40 who did not (no cancer outcome; NCO). Those samples were made up of 2 pairs of BO samples collected at two timepoints at separate locations within the oesophagus for all 80 patients (320 total). For 10 NCO patients, 2 biopsies at a third time point were also collected (20 additional for a total of 340). Barrett’s epithelium was isolated from underlying stroma as previously described (Li *et al*., 2015). For each patient a matched normal control (62 blood, 18 normal gastric tissue) was collected, and for 7 patients with blood controls available an additional matched control from adjacent normal tissue were also obtained. For full processing details, please refer to the original manuscript (Paulson *et al*., 2022).

Genomic DNA was extracted and sequenced at a median depth of 76X [range 40X - 106X] in the BO samples and 39X [range 29X - 64X] in the controls. Reads were aligned to the GRCh37 human reference genome using BurrowsWheeler Aligner (BWA). Somatic single-nucleotide variants (SNVs) were called using muTect v1.1.772 (Cibulskis *et al*., 2013), Strelka v1.0.1473 (Saunders *et al*., 2012), and LoFreq v2.1.3a74 (Wilm *et al*., 2012).

Somatic copy cumber aberrations (CNAs) were assessed in BO and control samples using the Omni 2.5 M 8v1.3 array and called using ASCAT (Van Loo *et al*., 2010). Mutational signatures were extracted and assigned using the SigProfiler framework (Alexandrov *et al*., 2020). All variant calls underwent stringent post-processing and quality control, detailed in the original manuscript.

For the current study, we directly used these high-confidence processed SNV and CNA calls as input for downstream analysis.

### Subclonal deconvolution using MOBSTER

We performed subclonal deconvolution on the 340 BO biopsies using MOBSTER (Caravagna, Heide, *et al*., 2020; Caravagna, Sanguinetti, *et al*., 2020), which allowed us to assign each SNV to either a clonal peak (pre- or post-amplification, where applicable), a subclonal expansion or the neutral tail of mutations that naturally arise during clonal expansion. Note that in the specific case of trisomy, the lower-VAF peak represents mutations of uncertain timing, as these cannot be unambiguously classified as pre- or post-amplification. The shape of the neutral tail allowed us to estimate the mutation rate per effective cell (Williams *et al*., 2016). Where adjacent CNA segments were within the 1 kbps and the ratios of the number of mutations on the pre-amplified and post-amplified peaks were similar, the CNA segments were merged. To ensure confident CNA timing downstream, we limited our subsequent analysis to CNAs with a total of 10 SNVs across the clonal peaks.

### Timing copy number alterations with Bayesian inference and evolution, CARBINE

We developed CARBINE, a hierarchical Bayesian method to infer the age of a patient at which specific copy number aberrations (CNAs) including genome doubling, trisomy and copy-number neutral LOH occurred in the history of a tumour. We applied a similar logic as previous research (Durinck *et al*., 2011; Greenman *et al*., 2012; Purdom *et al*., 2013; Cross *et al*., 2018; Gerstung *et al*., 2020), using the variant allele frequency (VAF) of single nucleotide variations (SNVs) to infer whether a given SNV occurred prior to or following the CNA. However, existing methods typically return the “molecular time” (i.e. the relative time prior to divergence) at which a CNA occurred. To translate this into the actual age of a patient at which a CNA occurs, we must account for the time since the most recent common ancestor (MRCA), which has previously only been possible with multi-region samples (Mitchell *et al*., 2018).

We have previously shown that the accumulation of effectively neutral passenger mutations results in a power-law-distributed “tail” in the VAF distribution (Williams *et al*., 2016), and that the shape of this tail encodes information about the “effective” mutation rate, *ν* (the average number of mutations per cell division divided by the fraction of successful cell divisions). In particular, we defined the cumulative number of mutations in the tumour, *M*(*f*), as a function of allelic frequency, *f*, as:

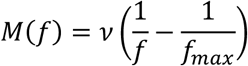

For CARBINE, we exploited this information to help infer the time since the MRCA for each patient. A hierarchical Bayesian approach was taken to ensure we extract the maximum amount of information from the data across multiple levels, whilst reducing the sample-specific parameters’ sensitivity to noise.

#### Parameters

Let there be *K* samples with each sample having *S_k_* distinct CNAs. The *k* subscript shall be used to refer to the *k^t^*^ℎ^ sample whilst the *i* subscript shall refer to the *i^t^*^ℎ^ CNA of the *k^t^*^ℎ^ sample.

Known Variables

- Number of SNVs present on the amplified and non-amplified alleles for the *i^t^*^ℎ^ CNA event in the *k^t^*^ℎ^sample, *α_k_*_,*i*_ and *β_k_*_,*i*_ respectively
- Number of SNVs that occurred on non-copy altered regions of DNA in the *k^t^*^ℎ^ sample, *γ_k_*
- Patient age at time of sampling, *T_k_*
- Size of the *i^t^*^ℎ^ copy-altered chromosome region in the *k^t^*^ℎ^ sample, *N_k_*_,*i*_ (in Mbp)
- Size of the non-copy altered regions of DNA in the *k^t^*^ℎ^ sample, *D_k_* (in Mbp; notably often 0 in genome doubled samples)
- The sample-specific “effective” mutation rate per division per Mbps, *υ_k_*

Sample-specific parameters

- Rate of aging-related mutations per allele per Mbp per year, *μ_k_*
- Age of the patient when the *i^t^*^ℎ^ CNA occurred (years), *t_k_*_,*i*_
- Time since sample MRCA (years), *τ_k_*
- The “effective” growth rate per year (fraction of cell divisions resulting in successful lineages multiplied by the cell division rate), *λ_k_*

Hyperparameters (population parameters)

- Mean of the population log(λk), ω
- Standard deviation of the population log(λk), σ

A number of assumptions were made in the development of this mathematical model. For completeness, they are listed below:

1. The patient specific SNV mutation rate (per base pair per year), *μ_k_*, of SNVs is approximately constant both across the genome and over the age of each patient (prior to the expansion of a single precancerous clone at the MRCA of the tumour).
2. Each patient’s net growth rate *λ_k_* is drawn from a lognormal population distribution.
3. The effective mutation rate per cell division, derived from the “tail” of the VAF distribution is equal to the mutation rate per year divided by the effective growth rate: 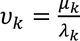

Given assumption (1), we assumed that the number of mutations that occur between the birth of the patient and the CNA follows a Poisson distribution with a shape parameter equal to the mutation rate per Mbp (*μ_k_*) multiplied by the genomic length of the change (*N_k_*_,*i*_), the age of the patient at the time of the CNA (*t_k_*_,*i*_) and the number of alleles contributing to the amplified peak (2 for tetraploidy and 1 for copy neutral LOH and trisomy). Similarly, the number of mutations that occurred following the CNA but prior to the MRCA will follow a Poisson distribution with a similar prefactor multiplied by the amount of time that passes between the CNA and the clonal expansion (*T_k_* − *τ_k_* − *t_k_*_,*i*_). We note a slight subtlety in the case of trisomy, upon the duplication of one of the chromosome arms, mutations that were present on the duplicated arm are at a frequency 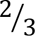, whilst mutations that were present on the non-duplicated arm are at a frequency 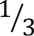, which is the frequency at which further mutations accumulate. This means that, unlike the LOH and tetraploidy case, where the number of SNVs on the amplified and non-amplified alleles directly correspond to the number of mutations that occurred before vs after the CNA, in trisomic regions the non-amplified peak *β_k_*_,*i*_ also contains SNVs that occurred prior to the amplification.

To provide additional information about the mutation rate, constraining the values of *τ_k_*, we assumed that the independently derived effective mutation rate per cell division, *υ_k_* is equal to the true mutation rate per year, *μ_k_*, multiplied by the effective growth rate, *λ_k_*. Furthermore, we placed a prior on the log-effective population size, *λ_k_ τ_k_*, constraining it to lie within realistic ranges (i.e. 95% of the probability mass lying between 10^2^ and 10^9^ cells). We similarly included a term to represent the number of mutations on non-copy altered regions which must have occurred in the time *T_k_* − *τ_k_*.

We ensured the positivity of the standard deviation of the population effective growth rate *σ* by truncating the normally distributed prior at 0 and the positivity of *λ_k_* and *μ_k_* naturally follows from the assumption that *λ_k_* is distributed log-normally and the effective mutation rate being strictly positive. *τ_k_* is constrained to lie between 0 and *T_k_* by placing a beta prior upon 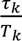, and *t_k,i_* is constrained to lie between 0 and *T_k_* − *τ_k_* by placing a beta prior upon 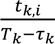.

In full, the model is as follows:

Priors

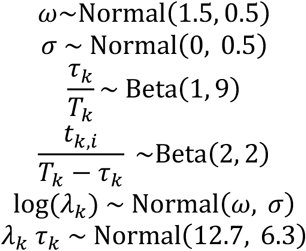

Parameter Transformations

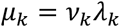

Likelihood

LOH

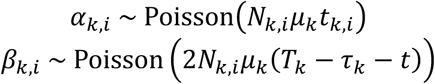

Tetraploid

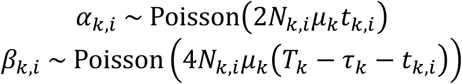

Trisomy

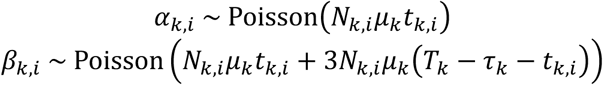

Diploid

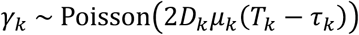

### Bayesian inference implementation

Bayesian inference was performed using Markov Chain Monte Carlo (MCMC). Specifically, the method was implemented in the probabilistic programming language Stan 3, a Hamiltonian Monte Carlo (HMC) variant that utilises the gradient of the posterior and automatically tunes sampling parameters to allow the sampler to efficiently explore the posterior (Carpenter *et al*., 2017). The inference was performed using the CmdStan (v2.34.1) interface and diagnostic statistics were calculated using the Arviz python package (v0.20.0) (Kumar *et al*., 2019).

A non-centred parameterisation of the hierarchical sampling statement (that is, 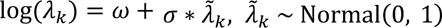) was used, which is mathematically equivalent to the centred version given above (log(*λ_k_*) ∼ Normal(*ω*, *σ*)) but ensures the sampler is able to explore areas of high curvature in the posterior which are common in hierarchical Bayesian models. This parameterization was chosen following the presence of divergent transitions (a Markov Chain transition in which the simulated Hamiltonian trajectory of the sampler path significantly differs from the true trajectory) in the centred parametrization that were not entirely eliminated by increasing target acceptance rate, which are indicative of a biased estimate of the posterior. Four independent Markov Chains with different initialisations (1000 warmup, 1000 sampling steps each) were run simultaneously and each converged to the same posterior, indicated by the sampler diagnostic Gelman-Rubin convergence statistic *R̂* < 1.1 for every parameter.

### Phylogenetic tree construction

The phylogenetic tree structure for patient 740 was derived from the framework reported previously (Paulson *et al*., 2022). Branch lengths were scaled according to the number of clonal single nucleotide variants (SNVs) located within diploid regions across all available samples.

For each somatic copy-number alteration (CNA) timed by CARBINE, we assessed its pattern of sharedness to classify the event as truncal (present in all samples), shared (present in a subset of samples), or private (restricted to a single sample). Using this classification, combined with the established phylogenetic structure, we assigned each CNA to the most parsimonious branch of the tree. A single real-time estimate for each CNA was then obtained by computing the median of the CARBINE-derived timings across all samples in which the event was present.

### Absolute timing of somatic copy number alterations in Barrett’s oesophagus

We next investigated how the rate of CNA acquisition varied over the clinically silent evolution to Barrett’s oesophagus. Across all patient samples, we first quantified the CNA accrual rate as the number of CNA events that occurred between the first CNA and the inferred MRCA time, where the posterior mean was used as a point estimate. Next, to assess whether the dynamics of chromosomal instability differed by clinical outcome, we compared the distribution of CNA rates between patients who progressed to oesophageal adenocarcinoma (cancer outcome, CO) and those who did not progress (no cancer outcome, NCO). Statistical differences were evaluated using both a Mann-Whitney U test (fig. 5A) and using a linear-mixed effects model, incorporating patient identity as a random effect to account for the inclusion of multiple CNAs from multiple biopsies within the same individual (fig. S2A).

Within each sample, we next evaluated if their dynamics of CNA acquisition better fitted a gradual or a punctuated model of CNA acquisition. We used a combination of the Kolmogorov-Smirnov test for uniformity of event times and a chi squared test to test whether the binned event times (with the number of bins equal to the total number of CNAs divided by 10, with a minimum of 2 bins) were significantly different from a uniform expectation. If either test was significant at 0.05, the sample was called as non-uniform.

### Evaluation of the accuracy of CARBINE estimates

We estimated the accuracy of the parameters inferred by CARBINE using i) simulated data, ii) clock-like SBS1 SNVs, iii) the multi-sampling WGS BO data, and iv) WGS data from OAC samples.

#### Simulated data

To evaluate the accuracy and robustness of CARBINE, we generated synthetic datasets modelling clonal evolution, somatic mutation accumulation, and copy number alterations (CNAs) using a custom simulation framework implemented in Python. This simulator captures the interplay between mutation timing, clonal growth dynamics, and sequencing noise, and was designed to reflect realistic features of cancer genome evolution.

We simulated data for K=300 virtual patients, each assigned an age uniformly sampled between 65 and 90 years. A subset (one-third) of patients underwent genome duplication (GD). For each patient, the number and location of CNAs were sampled based on autosomal chromosome sizes, with patients with GD acquiring the tetraploid state at a single random time point and non-GD patients receiving a random subset of 5–15 focal CNAs (LOH 50%, trisomy 40% and tetraploidy 10%) at randomly sampled times.

Growth rates were sampled from a log-normal distribution (μ=0.5, σ=0.3) to simulate inter-patient variability in clonal expansion. Mutation rates were sampled from a log-normal distribution centred at 0.03 mutations per Mb per year, with low dispersion (σ=0.001). For each CNA, we sampled a true mutation time using a truncated normal distribution centred on a patient-specific expected time since the MRCA, incorporating biological constraints such as maximum detectable clonal age and age at sampling.

We sampled from Poisson distributions to calculate the mutation counts both before and after each CNA event, assuming region-specific mutation rates and CNA-specific ploidy multipliers. Similarly, Poisson statistics were used to draw the number of clonal mutations present on the non-copy altered chromosomes. Additional passenger mutations were added to simulate the neutral tail. These mutations were drawn from a Pareto distribution consistent with neutral evolution.

To simulate sequencing noise, we modelled variant allele frequency (VAF) distributions using beta-binomial sampling for each category of mutation: clonal amplified, clonal non-amplified, and the neutral tail. Purities for each simulated sample were drawn from a beta distribution with shape parameters (45, 5) and the precision parameter for the beta-binomial noise was fixed at κ=80. Sequencing depth was sampled from a Poisson distribution with a mean of 100X (the same order as the ∼76X for the real data), with minimum coverage and read count thresholds applied to reflect realistic detection limits (e.g., minimum VAF = 0.05, minimum depth = 10X, and minimum alt reads = 3).

Each simulated sample yielded a set of SNVs and CNAs along with patient-specific metadata including age, purity, and effective mutation rate. These were processed using MOBSTER as above and CARBINE was run using identical parameters. The true CNA times and time since the MRCA were compared with the posterior median CARBINE results using linear regression.

#### Clock-like SBS1 point mutations

One of the key assumptions of CARBINE is that SNVs accrue in a linear fashion. To ensure that deviations from this assumption did not substantially bias the CNA timing, we re-ran CARBINE on just the set of clock-like SBS1 mutations, which are a well-validated mutational clock (Alexandrov *et al*., 2015). Mutational signature probabilities for each SNV were obtained from prior work (Paulson *et al*., 2022), in which SigProfiler was used to calculate the probability of each mutation belonging to SBS1, SBS2, SBS3, SBS5, SBS13, SBS17a, SBS17b, SBS18, SBS34 and SBS40 (Alexandrov *et al*., 2020). SNVs were designated as SBS1 mutations if that mutational signature probability was the maximum. SBS1 mutations represented 9.1% of all SNVs (485209 / 5350302).

We re-ran our Mobster subclonal deconvolution tool (Caravagna, Heide, *et al*., 2020; Caravagna, Sanguinetti, *et al*., 2020) on these reduced sets of SNVs, manually quality controlling every model fit. Due to the reduced number of SNVs, a number of CNA events now had fewer than the 10 SNV minimum cutoff and so were removed, leaving 908 timeable CNAs. Carbine was run on these SBS1 counts using the same priors and settings as above.

Comparison using all SNVs vs just SBS1 mutations was performed between the inferred CNA timings and the time since the MRCA using linear regression (p<2.2e-16 for both, R^2^=0.61 and R^2^=0.79 respectively).

#### Multi-sampling whole-genome sequencing data

Multi-region whole-genome sequencing allows for the identification of identical somatic copy-number alterations (CNAs) across spatially and/or temporally distinct biopsies from the same patient. We reasoned that, when two independent samples share an CNA (defined by concordant breakpoints and identical major and minor copy-number states), the alteration most likely arose prior to the emergence of the most recent common ancestor (MRCA) of the corresponding clones.

To assess the technical robustness of CARBINE’s CNA timing estimates, we first measured the absolute difference in inferred emergence time for shared CNAs across paired samples from the same patient. For each CNA, the real-time estimate was summarised as the median of its posterior distribution. We further examined the relationship between timing uncertainty (standard deviation of the posterior) and the degree of temporal discordance across samples, thereby evaluating whether posterior dispersion serves as an empirical proxy for confidence in CARBINE’s timing estimates.

To assess the biological plausibility of the inferred timings, CNAs were classified into three categories based on their clonality: truncal CNAs, shared across all samples from a patient; shared CNAs, present in a subset of samples; and private CNAs, detected in only one sample. Under the assumption that truncal alterations precede the MRCA of all samples, shared CNAs precede the MRCA of a subset of samples, and private CNAs occur after the MRCA of any sample pair, we tested whether CARBINE’s inferred timings reflect this expected temporal hierarchy. To control for inter-patient variability in age and progression dynamics, we used *relative time of occurrence* (defined as the fraction of time to the sample’s MRCA) for all comparisons. Statistical differences in the distribution of relative timings across the three CNA categories were assessed using a linear mixed-effects model, including the biopsy in which the CNA was timed as a random effect to account for potential clustering of CNAs within individual samples.

#### Validation of CNA timing in oesophageal adenocarcinoma samples

To validate that the features of CNA accrual that we observed in BO were also present in OAC, we downloaded the processed SNV and CNA data of the 97 primary OAC samples from the PCAWG cohort (Campbell *et al*., 2020). We used MOBSTER to perform subclonal deconvolution and peak assignment, restricting our analysis to those samples with high purity and which passed our manual QC step, leaving us with 38 samples, 20 of which were GD. We ran CARBINE on these data using the same priors and settings as our BO data, with all diagnostics suggesting that the MCMC chains had converged.

As with our BO data, we estimated the GD time by taking the median of all 2:1 and 2:2 segments. We took the age of the patient at the time of sampling as our estimate of OAC diagnosis time, as all our samples were derived from pre-treated primary tumours. The calculation of the CNA rate and detection of non-uniform CNA accrual was performed using the same process as for the BO samples above.

### Survival Analysis

Survival analyses were conducted to evaluate time to diagnosis of oesophageal adenocarcinoma (OAC), measured from the point of initial sampling. Only baseline (T1) samples were included. Samples exhibiting genome doubling were excluded due to their established association with increased risk of progression to OAC and their confounding impact on the temporal resolution of early copy-number events—whereby, for example, a simple, timable 2+1 alteration may be obscured after whole genome doubling by its transformation to a complex 4+2 copy number state.

After applying these filters, a single T1 sample was randomly selected per patient as the index lesion to avoid any intra-individual bias. Two genomic features – aneuploidy burden and somatic copy-number alteration (CNA) rate – were evaluated as prognostic covariates. Aneuploidy burden was defined as the total number of megabases affected by CNAs, as previously described by (Paulson *et al*., 2022). CNA rate was calculated by normalising the aneuploidy burden to the estimated time over which CNAs accrued, defined as the interval between the acquisition of the earliest CARBINE timable event and the most recent common ancestor (MRCA) of the index lesion.

To facilitate reproducible and unbiased dichotomisation of these continuous covariates, we applied the *maxstat.test* R function using the log-rank statistic, thereby identifying optimal cut-points for risk stratification. Univariate Cox proportional hazards models were fitted for each of the resulting binary covariates, and the concordance indices (C-indexes) were compared against each other using *compareC* R function, which implements a U-statistics-based approach for paired survival prediction comparisons. To further assess whether CNA rate provides prognostic information independent of overall aneuploidy burden, we also fitted a multivariate Cox model including both covariates. For this analysis, both variables were dichotomised at their respective medians to ensure balanced group sizes and comparable statistical power.

## Data Availability

Only reanalysis of previously published data was performed. The data from (Paulson *et al*., 2022) are available at the NCBI dbGaP database under accession code phs001912.v1.p1 [https://www.ncbi.nlm.nih.gov/projects/gap/cgi-bin/study.cgi?study_id=phs001912.v1.p1]. The open access data from (Alexandrov *et al*., 2020) are available from an Object Storage Bucket https://object.genomeinformatics.org in the icgc25k-open bucket.

## Code Availability

CARBINE software and code to reproduce the analysis accompany this submission and will be made publicly accessible via Github prior to publication.

## Author Contributions

This study was conceived and designed by CG and TAG. TAG funded and supervised the study. CG and TAG designed the Bayesian inference method, and were supported by SM, AS, WC and GC. The method was implemented and run by CG, and subsequent computational analysis was performed by CG and AFS, with support from HG and WC. The subclonal deconvolution tool was run by CG with support from SM, AS and GC. Data was provided by TGP and CM, and the results were interpreted by TGP, CM, NW, CG and TAG.

## Acknowledgements

This study was funded by Cancer Research UK (DRCNPG-May21_100001 to TAG), the Wellcome Trust (202778/Z/16/Z to TAG and AS), and the National Institute of Health (U54 CA217376 to TAG and CM). TAG and CG acknowledge support from the Thornton Foundation. CG acknowledges support from the Schmidt Fellows scheme at Imperial College London. A.F-S., is hosted by the Centro Nacional de Investigaciones Oncológicas (CNIO), which is supported by the Instituto de Salud Carlos III and recognized as a ‘Severo Ochoa’ Centre of Excellence (ref. CEX2019-000891-S) by the Spanish Ministry of Science and Innovation (MCIN/AEI/10.13039/501100011033). A.F-S. received the support of a fellowship from “La Caixa” Foundation (ID 100010434; LCF/BQ/DR21/11880009) and a mobility grant from the European Molecular Biology Organization (EMBO Scientific Exchange Grant 10898). CM acknowledges support from NIH grants U54 CA217376, U2C CA233254, R21 CA257980 and R01 CA140657 as well as CDMRP Breast Cancer Research Program Award BC132057.

