## Supplementary Information for "Protracted copy number alteration evolution in Barrett’s oesophagus leading to oesophageal cancer"

Gabbutt et.al.

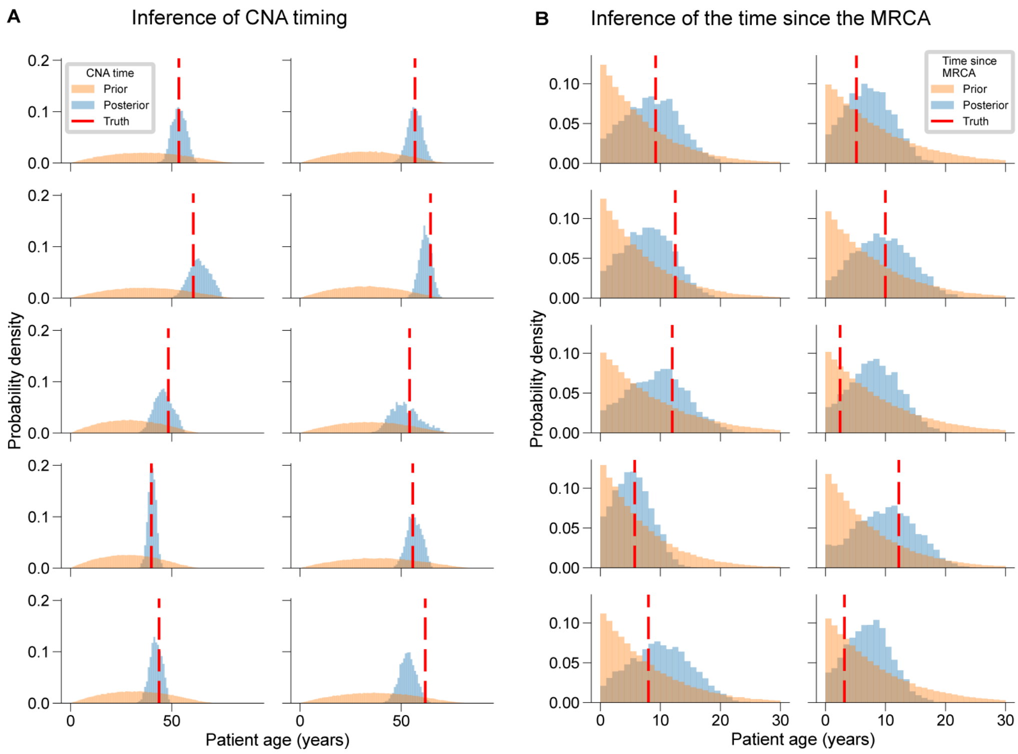


**Supplementary figure 1: Bayesian inference recovers accurate timings of CNAs and clonal expansions in simulated data**

**A:** Histograms showing the posterior distributions of CNA timing inferred by CARBINE for 10 representative simulated CNAs. Each panel shows the prior distribution (orange), posterior distribution (blue), and the true simulated CNA time (vertical red dashed line). In all cases, the posterior distributions exhibit strong shrinkage toward the true value, reflecting accurate recovery of event timing. **B:** Posterior distributions of the time since the most recent common ancestor (MRCA) for 10 representative samples. As in panel A, the prior (orange), posterior (blue), and true values (red dashed lines) are shown.

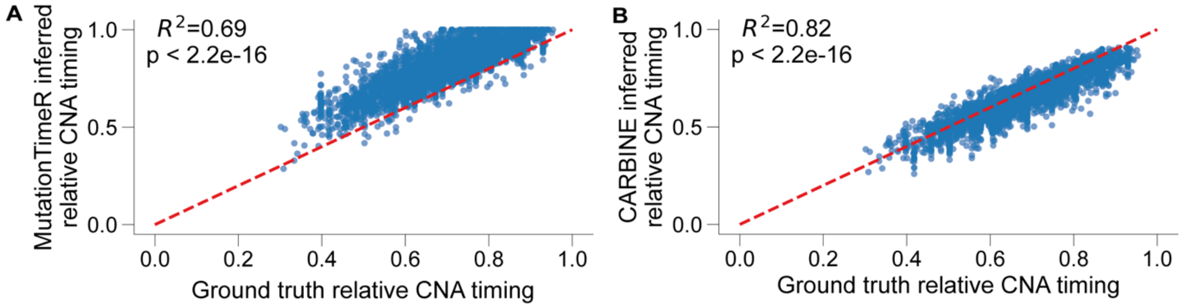


**Supplementary figure 2: CARBINE outperforms MutationTimeR at timing CNAs in simulated data**

Scatterplot of the ground truth simulated relative CNA time against the time inferred via MutationTimeR (**A**) and CARBINE (**B**), using the posterior mean as a summary statistic. The red dashed line corresponds to perfect agreement between the ground truth and inferred values. Only the 4223 CNA events that were successfully timed by both methods were included.

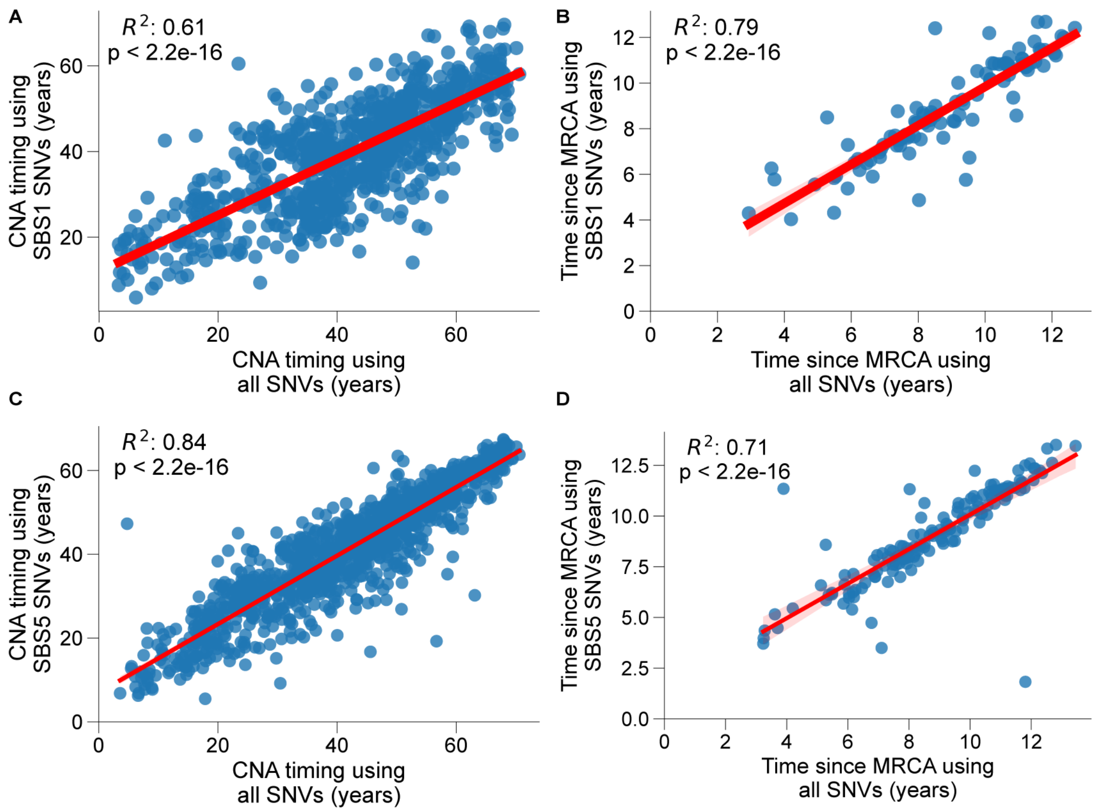


**Supplementary figure 3: Robustness of CNA timing in absolute time using clock-like SNVs**

Scatter plots comparing the inferred time at which a given CNA is inferred to have occurred (**A** & **C**) and the time since the most recent common ancestor (**B** & **D**) using all SNVs vs just those SNVs which likely belong to the aging signatures, SBS1 (**A**-**B**) and SBS5 (**C**-**D**). The least-squares best fit is overlaid in red ($p<2.2e-16$ in all cases, $R^2=0.61$, $R^2=0.79$, $R^2=0.84$ and $R^2=0.71$ respectively), together with bootstrapped 95% confidence intervals.

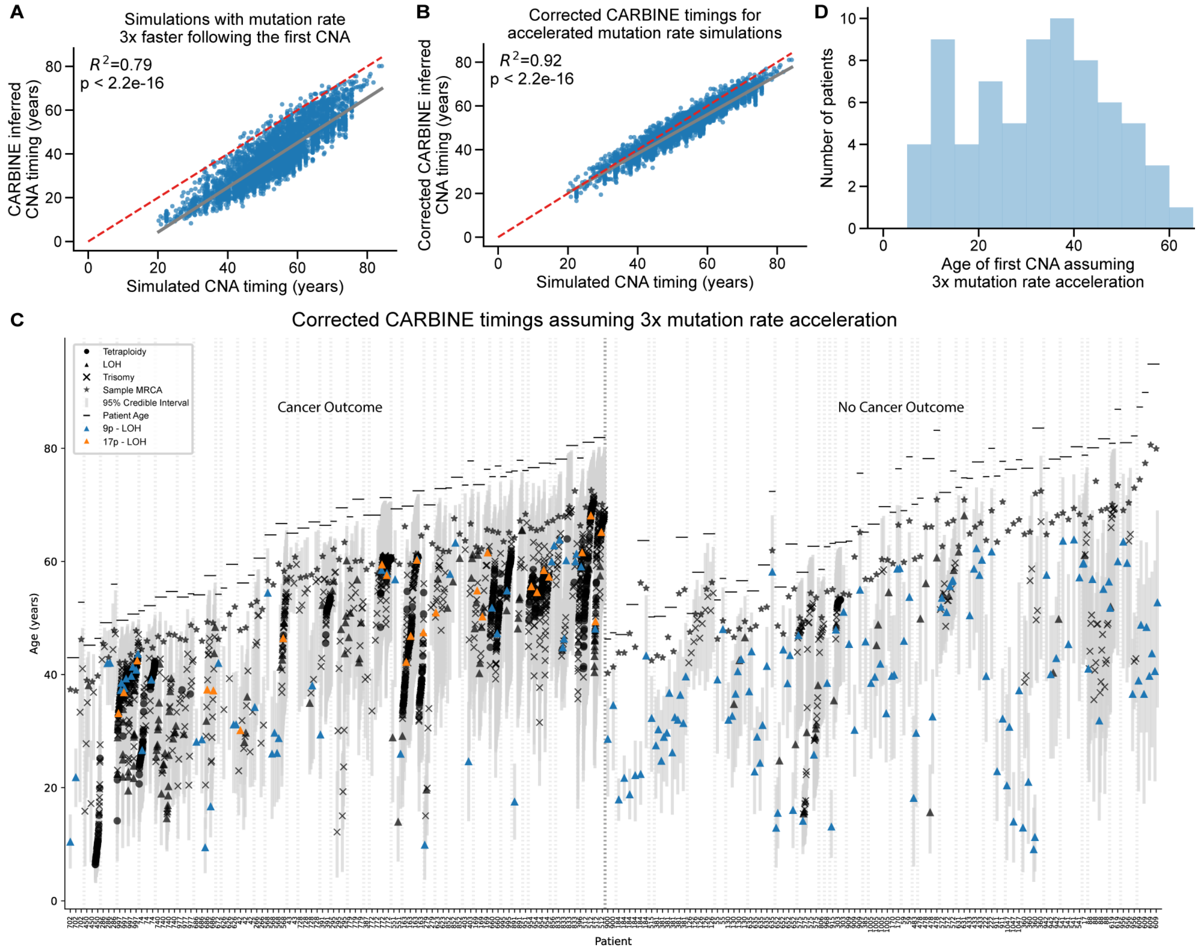


**Supplementary figure 4: CARBINE is robust to mutation rate acceleration**

**A-B:** Scatterplot of the simulated CNA time against the CARBINE inferred value (**A**) and against the CARBINE inferred value following correction (**B**), using the posterior mean as a summary statistic. The least squares fit is overlaid in grey with the 95% confidence interval calculated via bootstrapping. The red dashed line corresponds to perfect agreement between the ground truth and inferred values. **C:** Copy number alteration (CNA) timings for the 199 samples from 71 BO patients with timable CNAs, applying a correction assuming that the mutation rate increased 3-fold following the first CNA event. Each column represents one sample, and samples are grouped by patient. The posterior mean of individual CNA events are indicated by a shape (CN-LOH, triangle; trisomy, cross; tetraploidy, circle) and the 95% credible interval represented with a grey bar. The posterior mean of the MRCA is represented with a star and the patient age by a horizontal bar. Recurrent 9p LOH and 17p LOH events are annotated in blue or orange respectively. **D:** A histogram showing the age of the patient at the time of the first timable CNA event in any sample, assuming a 3x increase in the mutation rate following the first CNA event.

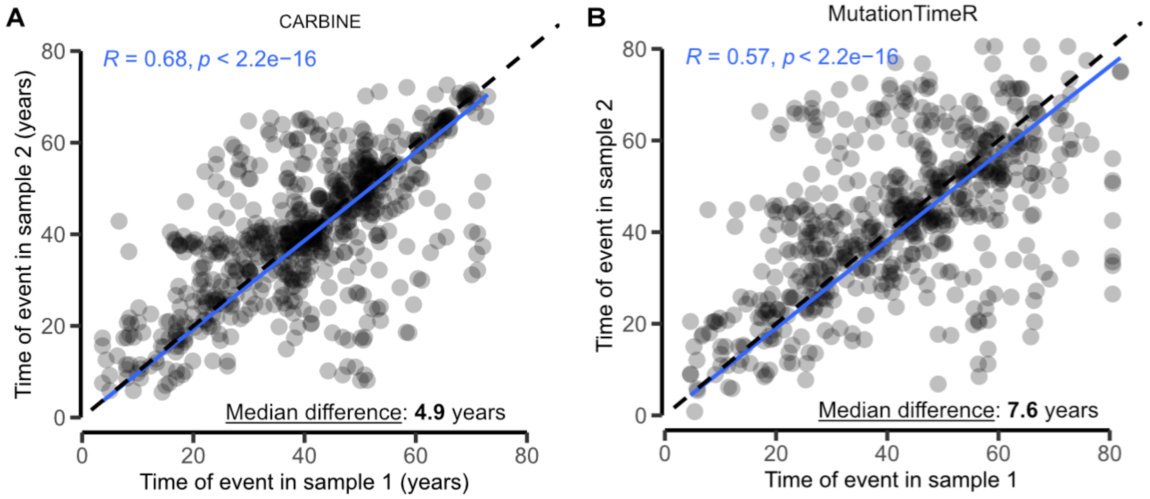


**Supplementary figure 5: Comparison of CARBINE and MutationTimeR CNA timing estimates using multiple samples**

**A-B:** Comparison of CARBINE (**A**) and MutationTimeR (**B**) timing estimates for identical somatic CNAs detected in pairs of spatially and/or temporally separated biopsies from the same patient. Each point represents the timing estimate for a single CNA in the first sample of the pair (x-axis) and the second sample (y-axis). The dashed diagonal line represents a perfect 1:1 correspondence. Spearman's correlation coefficient, statistical significance, and the best-fit linear regression are shown in blue. The median absolute differences in CNA timings are shown in the bottom right-hand corner (4.9y for CARBINE vs 7.6y for MutationTimeR).

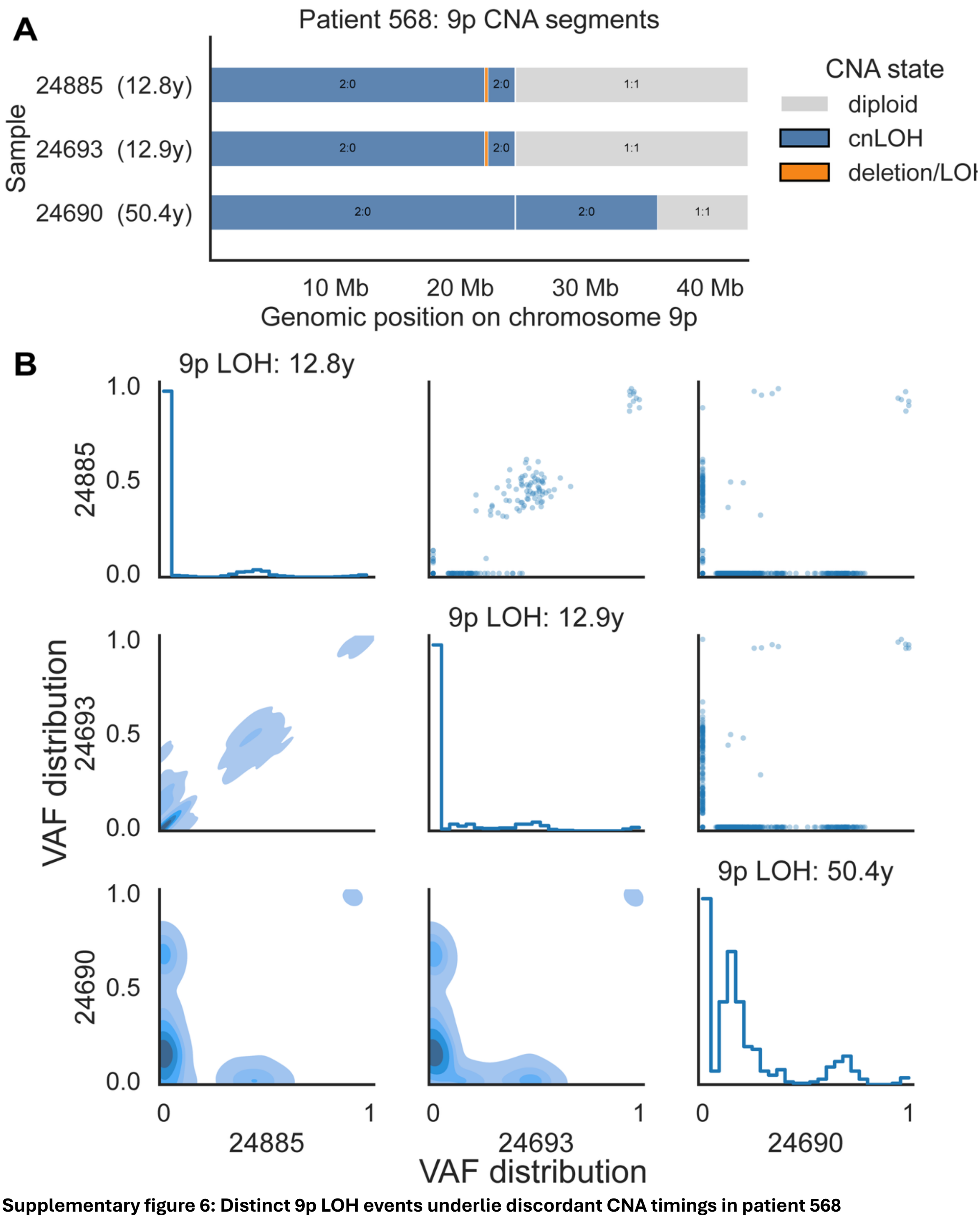


**Supplementary figure 6: Distinct 9p LOH events underlie discordant CNA timings in patient 568**
**A:** Allele-specific CNA segments across chromosome 9p for the three samples. Blue segments indicate copy-neutral LOH, orange indicates deletion/LOH, and grey indicates diploid regions. The late-timed sample 24690 has a distinct 9p LOH breakpoint from samples 24693 and 24885, supporting an independent LOH event. **B:** Pairwise comparison of variant allele frequencies (VAFs) for single nucleotide variants (SNVs) located within the common 9p LOH interval shared by the three samples. The diagonal panels show the marginal VAF distribution for each sample, the lower panels show kernel density estimates of the pairwise joint VAF distributions, and the upper panels show the corresponding scatter plots. CARBINE-inferred timings of the 9p LOH event are indicated above the diagonal panels for each sample.

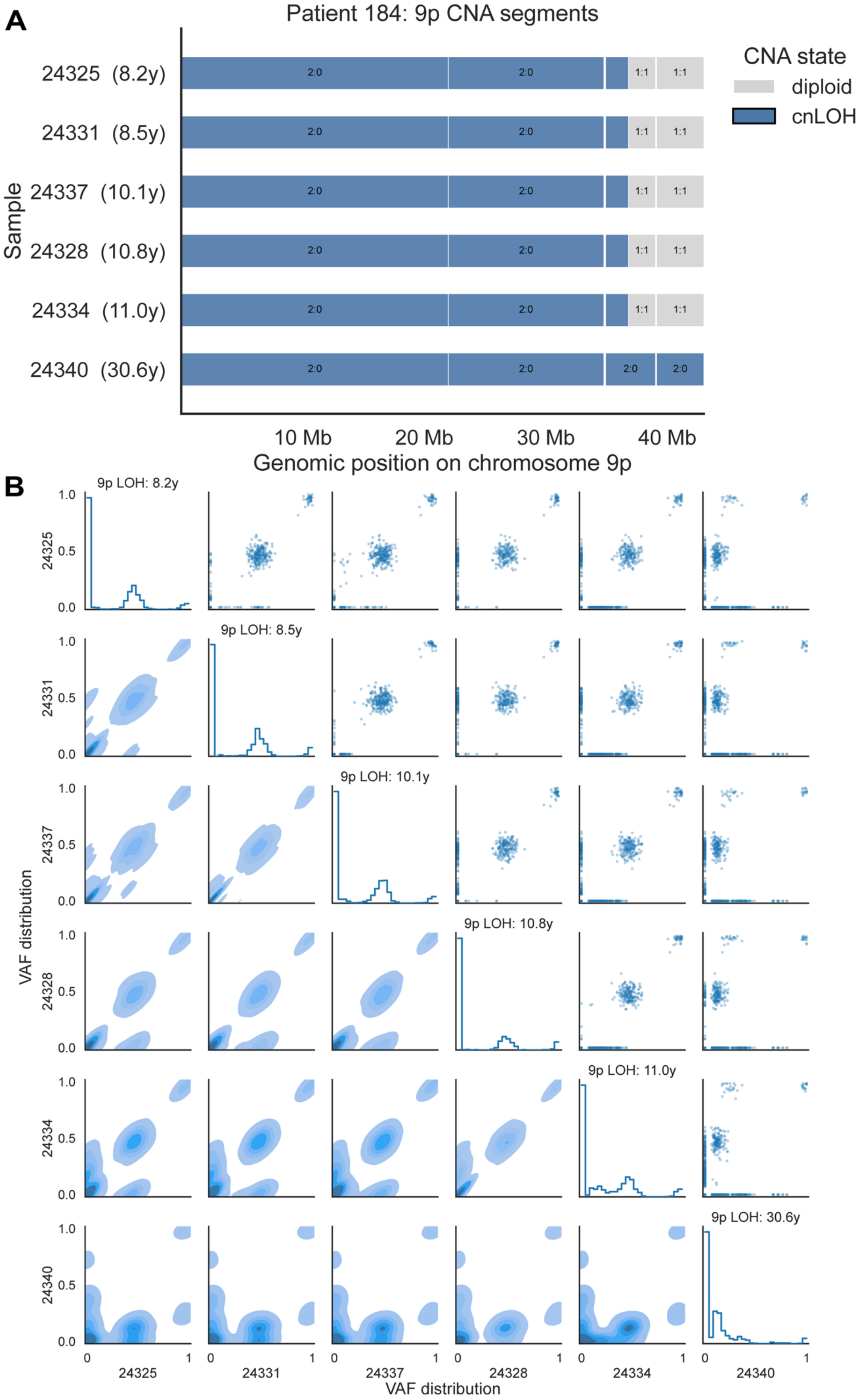
A
Patient 184: 9p CNA segments
CNA state
diploid
cnLOH
Sample
24325 (8.2y)
24331 (8.5y)
24337 (10.1y)
24328 (10.8y)
24334 (11.0y)
24340 (30.6y)
2:0
1:1
10 Mb
20 Mb
30 Mb
40 Mb
Genomic position on chromosome 9p
B
9p LOH: 8.2y
9p LOH: 8.5y
9p LOH: 10.1y
9p LOH: 10.8y
9p LOH: 11.0y
9p LOH: 30.6y
VAF distribution
24325
24331
24337
24328
24334
24340
1.0
0.5
0.0
0
1
VAF distribution

**Supplementary figure 7: Distinct 9p LOH events underlie discordant CNA timings in patient 184**
**A:** Allele-specific CNA segments across chromosome 9p for six samples. Blue segments indicate copy-neutral LOH and grey segments indicate diploid regions. Samples 24325, 24331, 24337, 24328 and 24334 show the same 9p cnLOH segment boundaries, whereas sample 24340 shows a broader 9p cnLOH segment. **B:** Pairwise comparison of variant allele frequencies (VAFs) for single nucleotide variants (SNVs) located within the common 9p LOH interval shared by the three samples. The diagonal panels show the marginal VAF distribution for each sample, the lower panels show kernel density estimates of the pairwise joint VAF distributions, and the upper panels show the corresponding scatter plots. CARBINE-inferred timings of the 9p LOH event are indicated above the diagonal panels for each sample. Together, these patterns are consistent with sample 24340 being dominated by a distantly related clone that acquired a later, broader 9p cnLOH event, with a minor contribution from the early 9p cnLOH clone present in the other five samples.

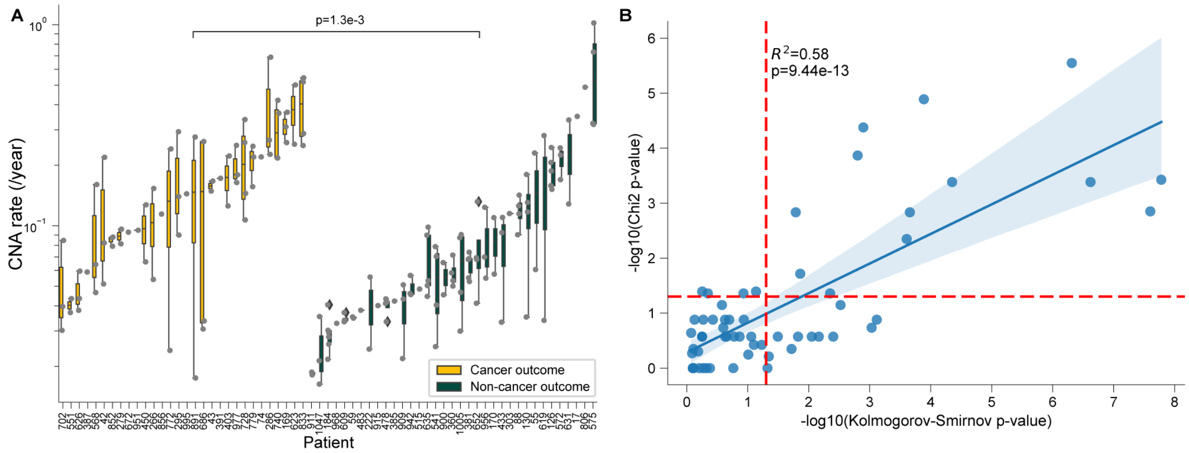


**Supplementary figure 8: Concordance between statistical tests for detecting punctuated CNA acquisition**
**A:** Boxplot comparing the rate of copy number alteration (CNA) acquisition per year in patients with (CO) and without (NCO) cancer outcomes, excluding those with genomic doubling (GD) events. Each point represents an individual patient's estimated CNA rate derived from the timing of somatic mutations within CNA segments. Patients with cancer outcomes exhibit significantly higher CNA accumulation rates than those without (linear mixed model, p=1.3e-3). **B:** Scatter plot comparing results from two statistical methods used to distinguish punctuated versus uniform CNA acquisition across samples. Each point represents a sample, plotted by the –$\log_{10}$-transformed Kolmogorov–Smirnov (x-axis) and Chi-squared (y-axis) test p-values. Both tests compare the observed distribution of CNA times to those expected under a uniform accumulation model. Red dashed lines represent a significance threshold of p = 0.05. A significant positive correlation is observed between the two test statistics (p=9.4e-13, $R^2$=0.58).

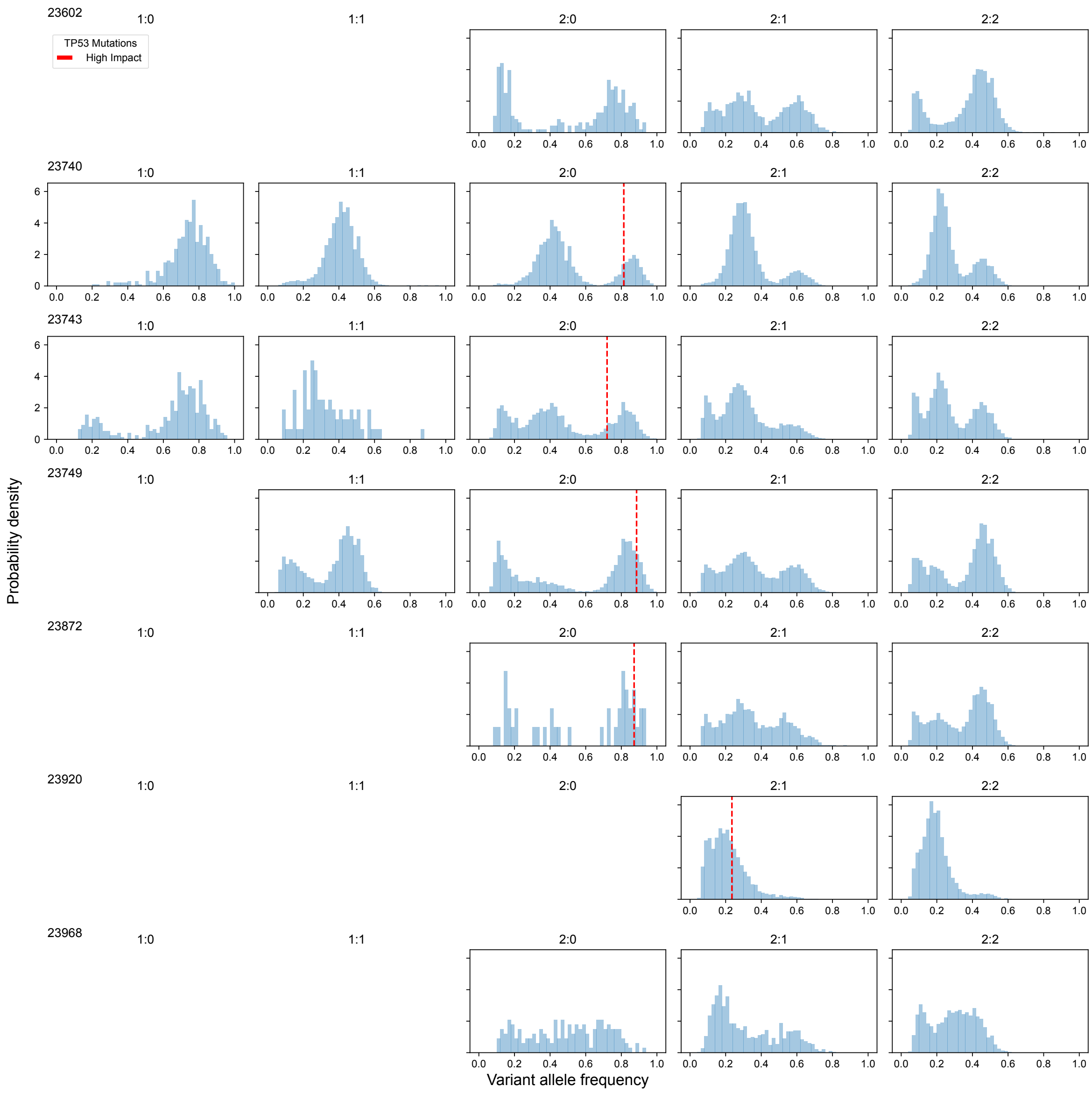


**Supplementary figure 9: Relationship between TP53 inactivation and WGD.**
Histograms showing the variant allele frequency distributions (VAF), separated by the different copy states (1:0 Loss, 1:1 Diploid, 2:0 Copy neutral loss of heterozygosity, 2:1 Trisomy and 2:2 tetraploidy). Each row is a different sample. TP53 mutations are overlaid as vertical dashed red lines centred on the VAF of the SNV. All non-synonymous TP53 mutations in these samples were annotated as high impact.

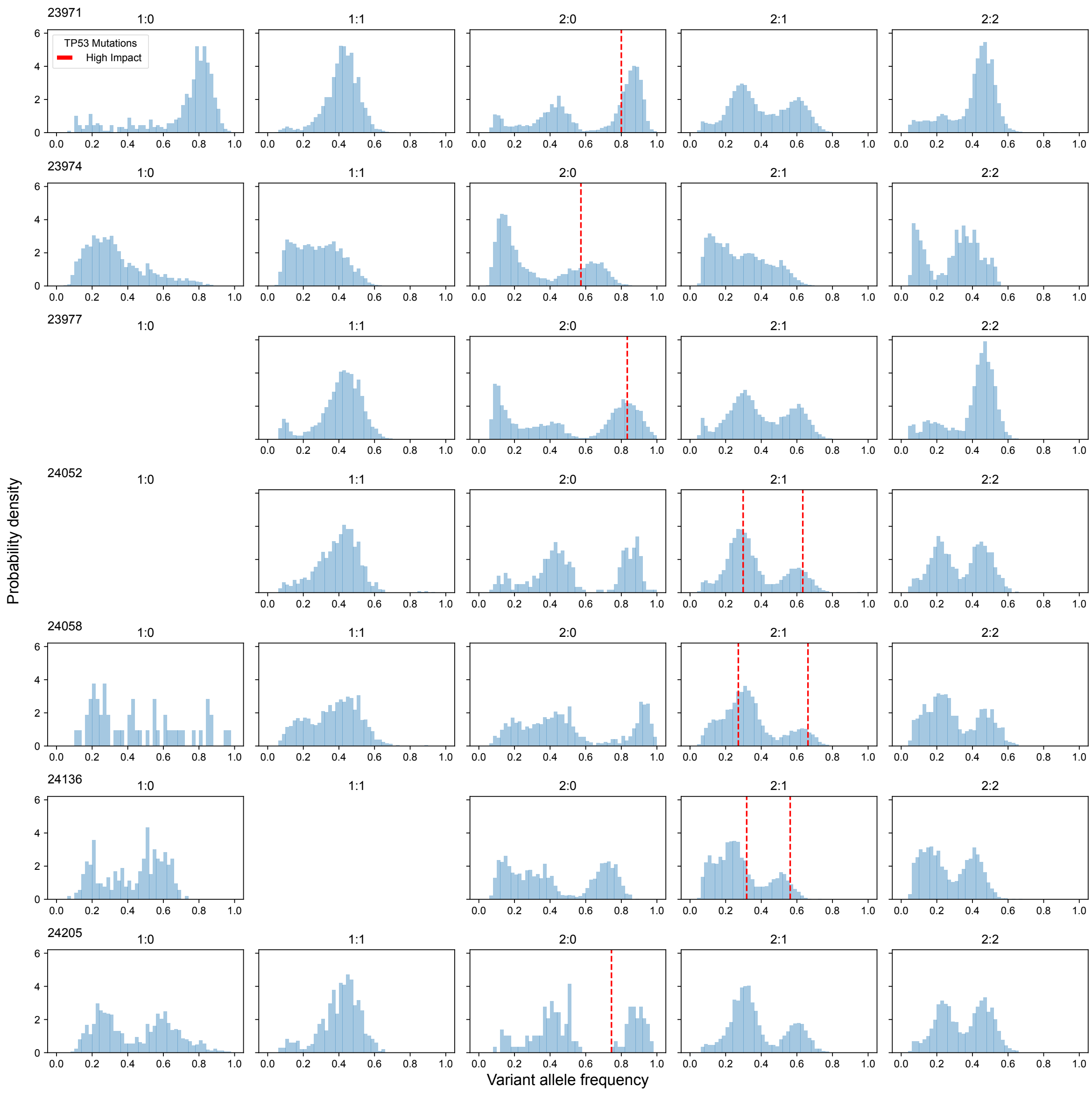


**Supplementary figure 10: Relationship between TP53 inactivation and WGD.**
Histograms showing the variant allele frequency distributions (VAF), separated by the different copy states (1:0 Loss, 1:1 Diploid, 2:0 Copy neutral loss of heterozygosity, 2:1 Trisomy and 2:2 tetraploidy). Each row is a different sample. TP53 mutations are overlaid as vertical dashed red lines centred on the VAF of the SNV. All non-synonymous TP53 mutations in these samples were annotated as high impact.

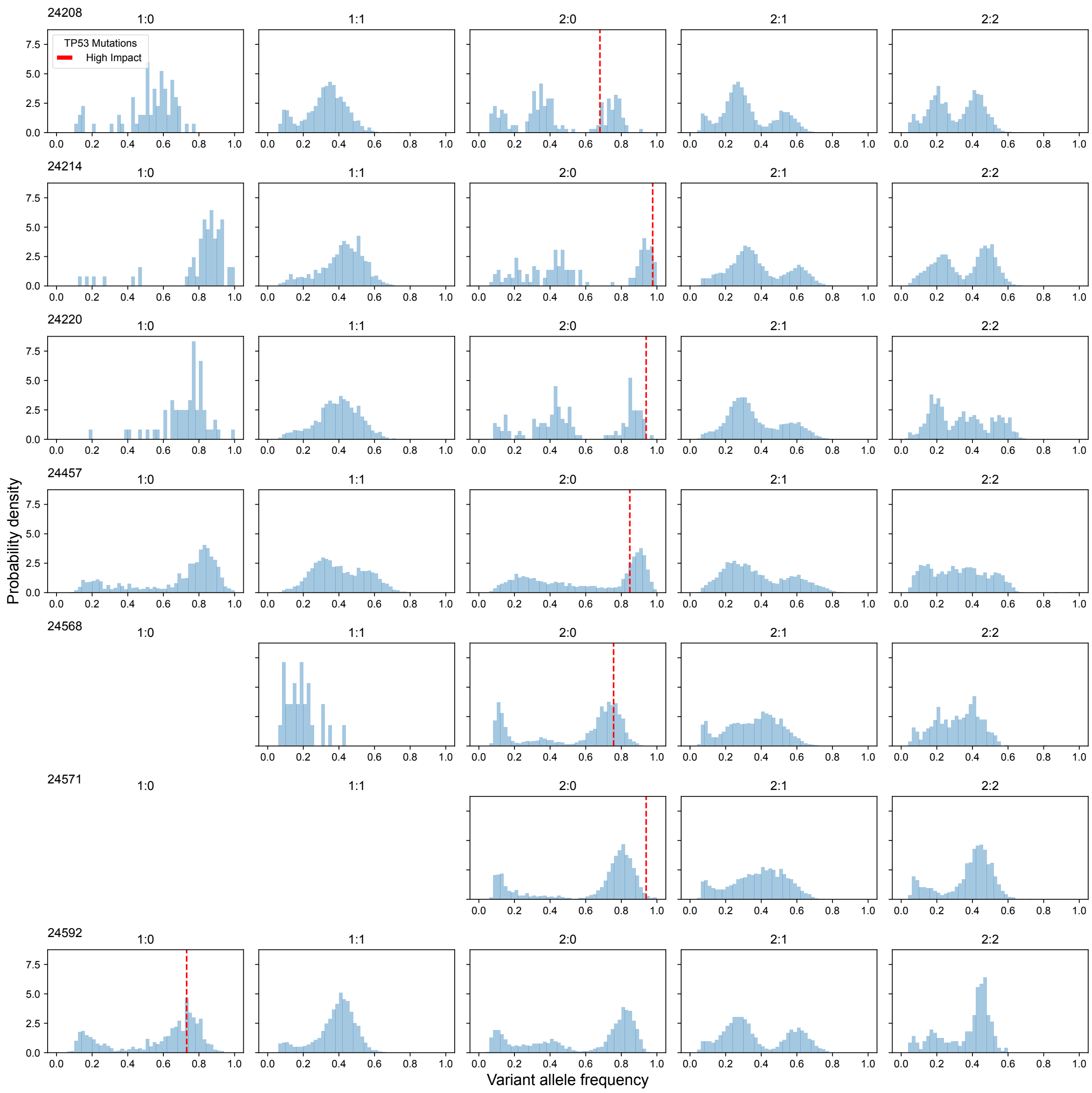


**Supplementary figure 11: Relationship between TP53 inactivation and WGD.**
Histograms showing the variant allele frequency distributions (VAF), separated by the different copy states (1:0 Loss, 1:1 Diploid, 2:0 Copy neutral loss of heterozygosity, 2:1 Trisomy and 2:2 tetraploidy). Each row is a different sample. TP53 mutations are overlaid as vertical dashed red lines centred on the VAF of the SNV. All non-synonymous TP53 mutations in these samples were annotated as high impact.

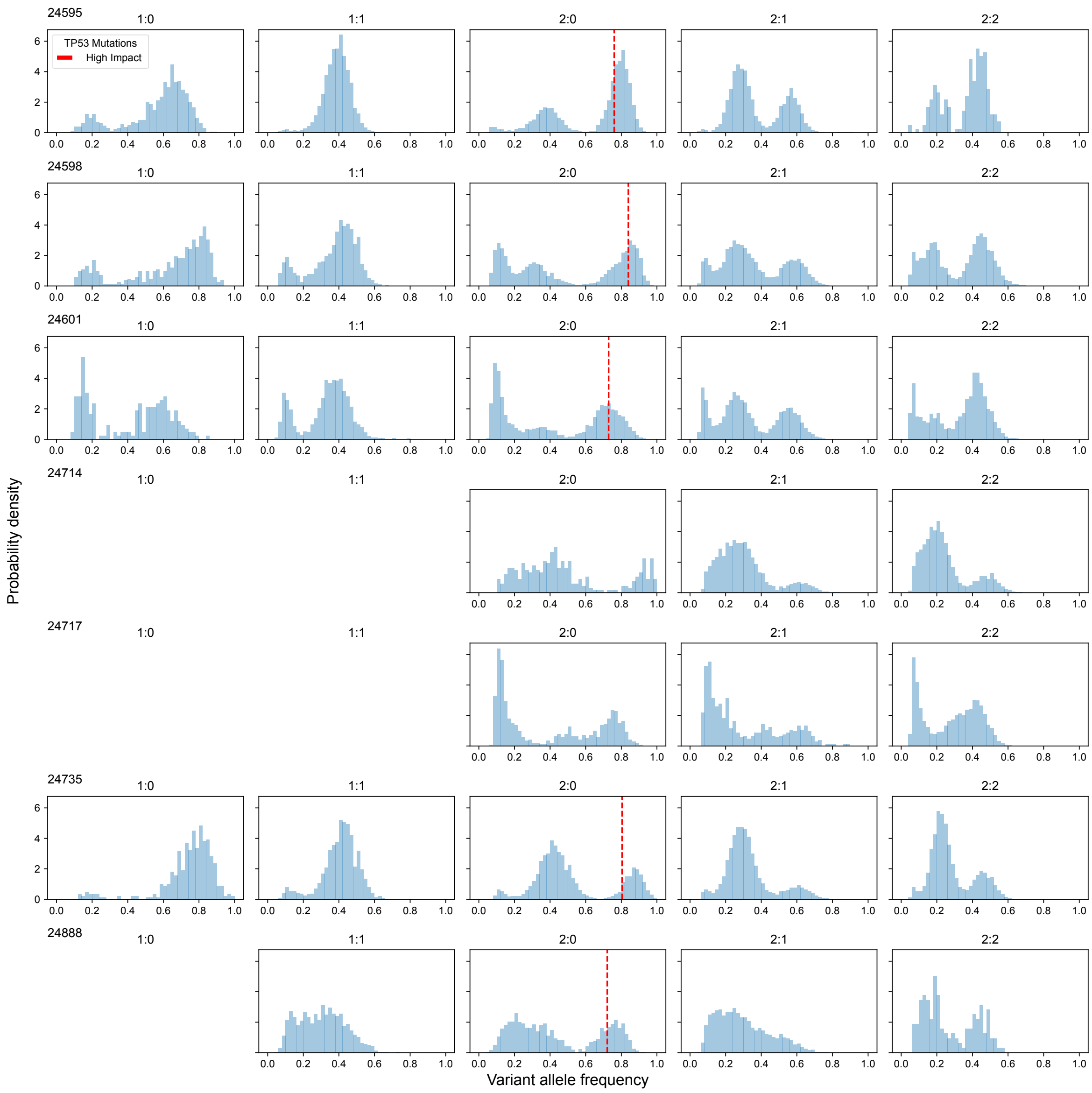


**Supplementary figure 12: Relationship between TP53 inactivation and WGD.**
Histograms showing the variant allele frequency distributions (VAF), separated by the different copy states (1:0 Loss, 1:1 Diploid, 2:0 Copy neutral loss of heterozygosity, 2:1 Trisomy and 2:2 tetraploidy). Each row is a different sample. TP53 mutations are overlaid as vertical dashed red lines centred on the VAF of the SNV. All non-synonymous TP53 mutations in these samples were annotated as high impact.

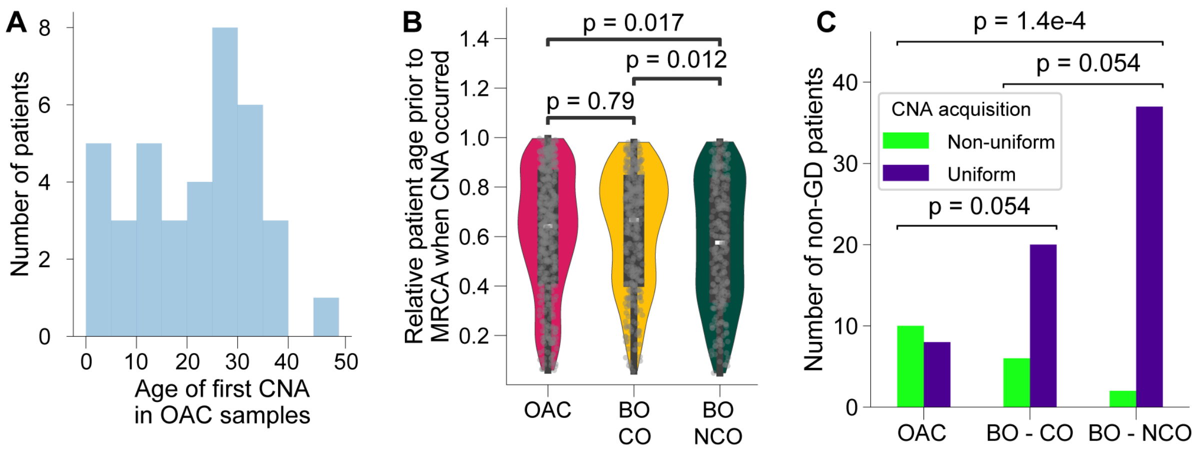


**Supplementary figure 13: CNA timing in oesophageal adenocarcinoma data.**
**A:** Histogram of the age of the OAC patient when the first CNA occurred in years. **B:** Violin plots showing the relative CNA timing ($\frac{t_{CNA}}{T-\tau}$) between PCAWG OAC samples, Barrett's oesophagus patient with a cancer outcome and no cancer outcome. Violin plot: centre line, median; box limits, upper and lower quartiles; whiskers, 1.5x interquartile range; points, outlier; p values, Mann-Whitney U tests - Holm-Bonferroni correction. **C:** Bar plot showing the number of patients without WGD with at least 1 sample with a non-uniform pattern of CNA acquisition vs all uniform samples is higher in progressors than non-progressors (pairwise Fisher's exact tests, Benjamini-Hochberg corrected).

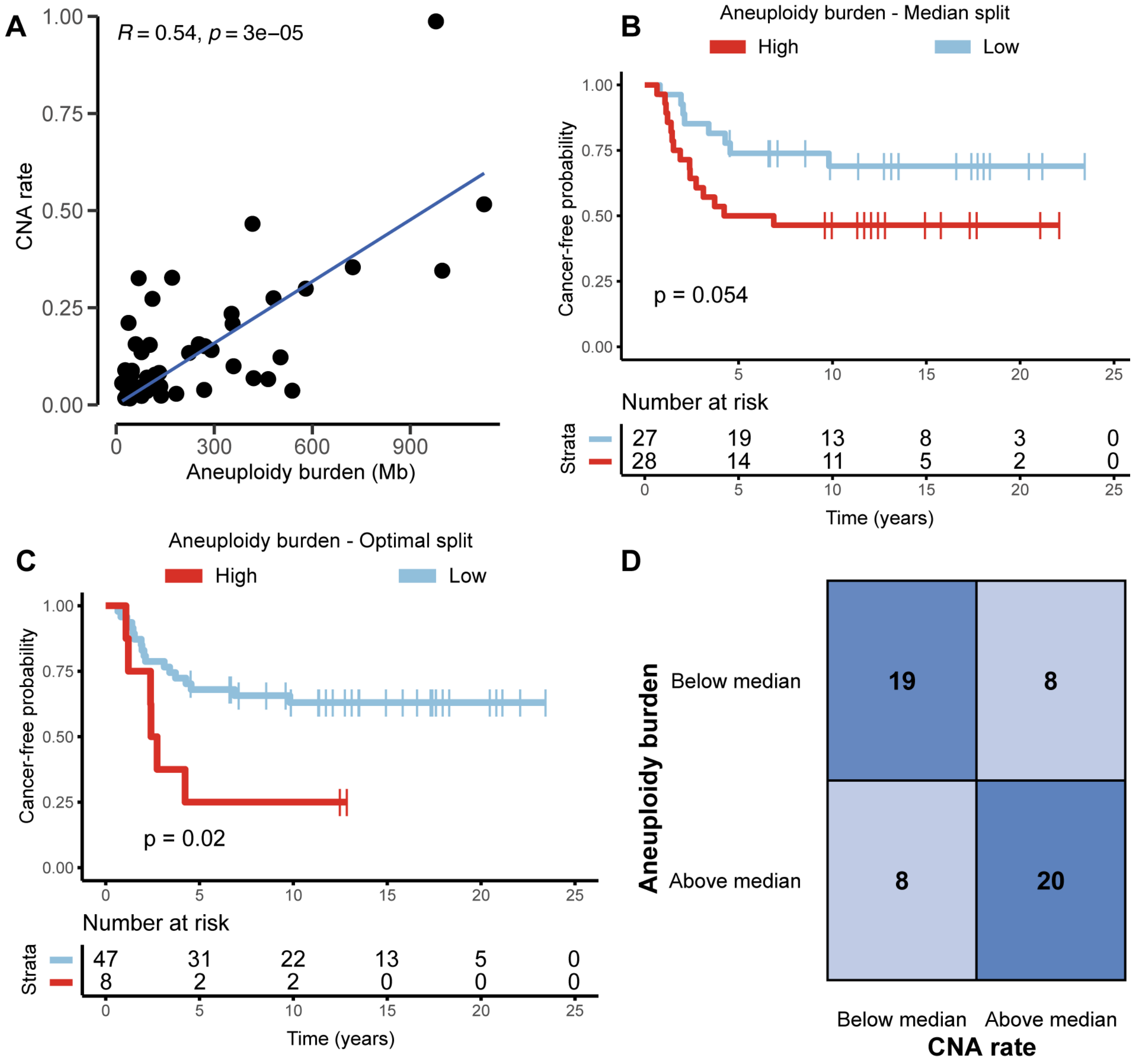


**Supplementary figure 14: Comparison of CNA burden and CNA rate as prognostic markers of progression to oesophageal cancer**

**A:** Scatterplot showing the relationship between the rate of CNA acquisition and the total somatic CNA burden in non-WGD samples. **B-C:** Kaplan–Meier analyses of time to progression to oesophageal adenocarcinoma in patients stratified by high (red) or low (blue) aneuploidy burden. Aneuploidy burden was quantified as the fraction of genome altered (FGA), defined as the proportion of megabases affected by somatic copy number alterations, as described previously (Paulson et al. 2022). P-value was estimated using a log-rank test, and the threshold for high and low FGA were determined by median split (**B**) or by optimizing the log-rank test statistic (**C**). **D:** Confusion matrix illustrating the discordance in patient stratification when using aneuploidy burden (y-axis) versus the rate of CNA acquisition (x-axis) as biomarkers. The thresholds applied to both numerical metrics were their respective median cohort values.

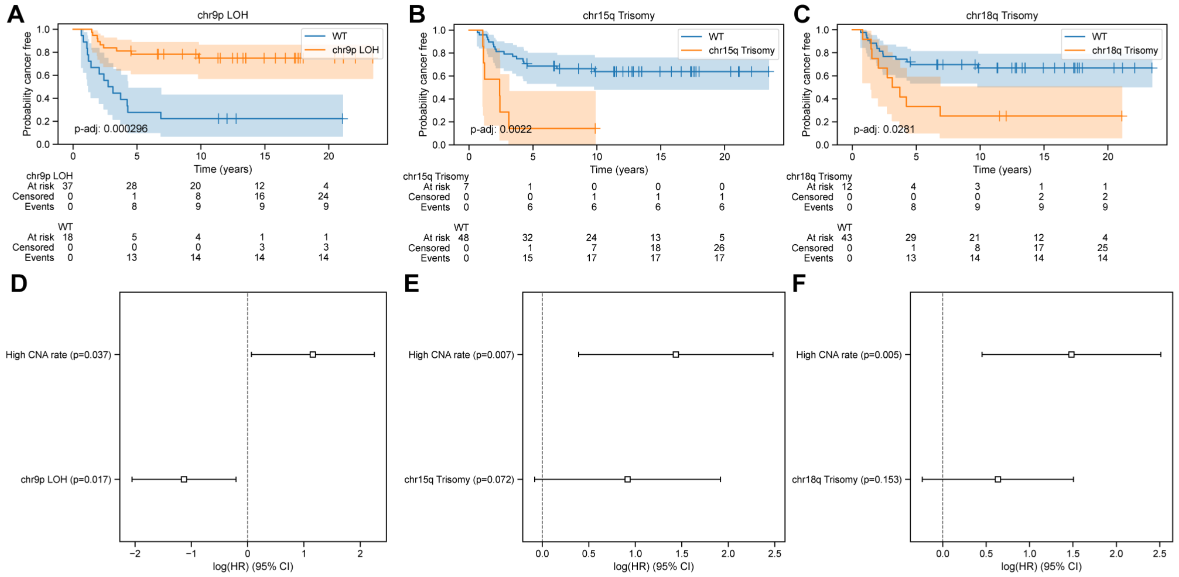


**Supplementary figure 15: Testing the prognostic potential of recurrent CNAs**

**A-C:** Kaplan–Meier analyses of time to progression to oesophageal adenocarcinoma in patients stratified by the presence or absence of 9p loss of heterozygosity (LOH, **A**), 15q trisomy (**B**) and 18q trisomy (**C**). P-values were estimated using a log-rank test and were multiple test corrected using the Benjamini/Hochberg method. **D-E:** Forest plots showing the log hazard ratios (HR) for a multivariate cox regression including the CNA rate and the presence of 9p LOH (**D**), 15q trisomy (**E**) and 18q trisomy (**F**).

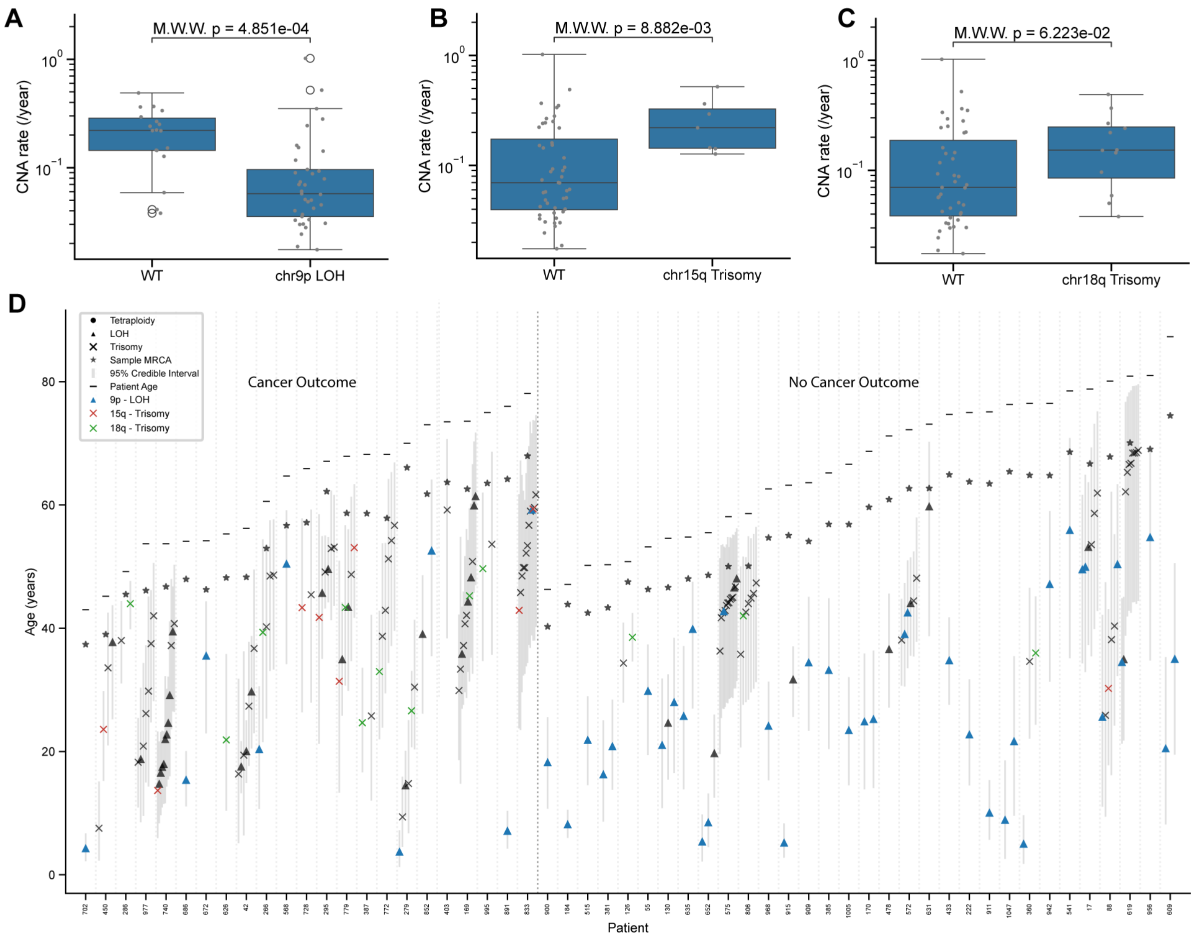


**Supplementary figure 16: Recurrent CNAs are associated with altered CNA accrual rates**

**A-C:** Box plots showing the CNA acquisition rates per year separated by the presence or absence of 9p LOH (**A**), 15q trisomy (**B**) and 18q trisomy (**C**). p values were calculated using a Mann Whitney U test. **D:** CNA timings for the 55 highest SCA T1 samples from BO patients without GD. Each column represents one sample. The posterior mean of individual CNA events are indicated by a shape (CN-LOH, triangle; trisomy, cross; tetraploidy, circle) and the 95% credible interval represented with a grey bar. The posterior mean of the MRCA is represented with a star and the patient age by a horizontal bar. Recurrent 9p LOH and 15q trisomy and 18q trisomy events are annotated in blue, red or green respectively.

### Supplementary Methods

#### Impact of acceleration in the mutation rate

To evaluate the impact of the mutation rate on CNA timing, we derived an analytic correction to the inferred timing, validated this correction using simulated data and applied it to our real data (supplementary fig. 4).

Derivation of an analytic correction accounting for mutation rate acceleration

Consider a system with the same setup and notation as in the manuscript, except prior to the first CNA, point mutations are accrued at a rate $\rho\mu_k$ per allele per Mbp per year vs a mutation rate of $\mu_k$ after the first CNA event (i.e. $\rho = 0.5$ represents a 2x acceleration in mutation rates across the genome following the first CNA).

We shall use a moment matching approach to derive this analytic correction factor of the copy number timings. First, we shall consider the system with constant mutation rate, as in the manuscript.

Likelihood

LOH

$$\alpha_{k,i} \sim \text{Poisson}\left(N_{k,i}\mu_k t_{k,i}\right) \quad \text{S1}$$

$$\beta_{k,i} \sim \text{Poisson}\left(2N_{k,i}\mu_k\left(T_k - \tau_k - t_{k,i}\right)\right) \quad \text{S2}$$

Tetraploid

$$\alpha_{k,i} \sim \text{Poisson}\left(2N_{k,i}\mu_k t_{k,i}\right) \quad \text{S3}$$

$$\beta_{k,i} \sim \text{Poisson}\left(4N_{k,i}\mu_k\left(T_k - \tau_k - t_{k,i}\right)\right) \quad \text{S4}$$

Trisomy

$$\alpha_{k,i} \sim \text{Poisson}\left(N_{k,i}\mu_k t_{k,i}\right) \quad \text{S5}$$

$$\beta_{k,i} \sim \text{Poisson}\left(N_{k,i}\mu_k t_{k,i} + 3N_{k,i}\mu_k\left(T_k - \tau_k - t_{k,i}\right)\right) \quad \text{S6}$$

We can equate the observed numbers of SNVs observed on each peak with the expectation of the Poisson distribution at our CNA timing estimate $t_{k,i}^{est}$, as in previous work (Durinck *et al.*, 2011; Greenman *et al.*, 2012; Purdom *et al.*, 2013; Cross *et al.*, 2018; Gerstung *et al.*, 2020):

LOH

$$\alpha_{k,i} = N_{k,i}\mu_k t_{k,i}^{est} \quad \text{S7}$$

$$\beta_{k,i} = 2N_{k,i}\mu_k\left(T_k - \tau_k - t_{k,i}^{est}\right) \quad \text{S8}$$

Tetraploid

$$\alpha_{k,i} = 2N_{k,i}\mu_k t_{k,i}^{est} \quad \text{S9}$$

$$\beta_{k,i} = 4N_{k,i}\mu_k\left(T_k - \tau_k - t_{k,i}^{est}\right) \quad \text{S10}$$

Trisomy

$$\alpha_{k,i} = N_{k,i}\mu_k t_{k,i}^{est} \quad \text{S11}$$

$$\beta_{k,i} = N_{k,i}\mu_k t_{k,i}^{est} + 3N_{k,i}\mu_k\left(T_k - \tau_k - t_{k,i}^{est}\right) \quad \text{S12}$$

Combining these pairs of equations (e.g. S7 and S8 to give S13) yields a point estimate of the CNA timing in molecular time under a constant mutation rate, $\tilde{t}_{k,i} = \frac{t_{k,i}}{T_k - \tau_k}$, only depending on $\alpha_{k,i}$ and $\beta_{k,i}$.

LOH

$$\tilde{t}_{k,i}^{est} = \frac{2\alpha_{k,i}}{2\alpha_{k,i} + \beta_{k,i}} \qquad \text{S13}$$

Tetraploid

$$\tilde{t}_{k,i}^{est} = \frac{2\alpha_{k,i}}{2\alpha_{k,i} + \beta_{k,i}} \qquad \text{S14}$$

Trisomy

$$\tilde{t}_{k,i}^{est} = \frac{3\alpha_{k,i}}{2\alpha_{k,i} + \beta_{k,i}} \qquad \text{S15}$$

Let us now consider the true timing of the first CNA, which we shall denote without loss of generality as $\tilde{t}_{k,i=1}^{true}$, in the case where we allow the mutation rate prior to the CNA occurring to be $\rho\mu_k$. Following an identical logic as above, the moment matching formulae become:

LOH

$$\alpha_{k,i} = \rho N_{k,i}\mu_k t_{k,1}^{true} \qquad \text{S16}$$

$$\beta_{k,i} = 2N_{k,i}\mu_k\left(T_k - \tau_k - t_{k,1}^{est}\right) \qquad \text{S17}$$

Tetraploid

$$\alpha_{k,i} = 2\rho N_{k,i}\mu_k t_{k,i}^{true} \qquad \text{S18}$$

$$\beta_{k,i} = 4N_{k,i}\mu_k\left(T_k - \tau_k - t_{k,1}^{true}\right) \qquad \text{S19}$$

Trisomy

$$\alpha_{k,i} = \rho N_{k,i}\mu_k t_{k,1}^{true} \qquad \text{S20}$$

$$\beta_{k,i} = \rho N_{k,i}\mu_k t_{k,1}^{true} + 3N_{k,i}\mu_k\left(T_k - \tau_k - t_{k,1}^{true}\right) \qquad \text{S21}$$

Which can be combined as above (e.g. S16 and S17 to give S22) to yield the expression for the true first CNA timing in molecular time in terms of $\alpha_{k,i}$, $\beta_{k,i}$ and $\rho$:

LOH

$$\tilde{t}_{k,1}^{true} = \frac{2\alpha_{k,1}}{2\alpha_{k,1} + \rho\beta_{k,1}} \qquad \text{S22}$$

Tetraploid

$$\tilde{t}_{k,1}^{true} = \frac{2\alpha_{k,1}}{2\alpha_{k,1} + \rho\beta_{k,1}} \qquad \text{S23}$$

Trisomy

$$\tilde{t}_{k,1}^{true} = \frac{3\alpha_{k,1}}{(3-\rho)\alpha_{k,1} + \rho\beta_{k,1}} \qquad \text{S24}$$

We can thus combine these pairs of equations (e.g. S13 and S22) to calculate the true molecular CNA timing purely in terms of the constant mutation rate timing estimate and $\rho$. All three cases reduce to the same expression for $\tilde{t}_{k,1}^{true}$:

$$\tilde{t}_{k,1}^{true} = \frac{\tilde{t}_{k,1}^{est}}{\rho + (1-\rho)\tilde{t}_{k,1}^{est}} \qquad \text{S25}$$

This expression holds for the first CNA event. For subsequent CNA events the same logic holds, except only the interval before the first CNA is affected by the reduced mutation rate. Because mutations accrue at rate $\rho\mu_k$ before the first CNA and at rate $\mu_k$ afterwards, the total expected number of mutations accumulated along a single ancestral lineage up to event $i$ is proportional to:

$$\rho\tilde{t}_{k,1}^{true} + \left(\tilde{t}_{k,i}^{true} - \rho\tilde{t}_{k,1}^{true}\right) \qquad \text{S26}$$

Similarly, the total expected number accumulated over the full interval is proportional to:

$$\rho\tilde{t}_{k,1}^{true} + \left(1 - \tilde{t}_{k,1}^{true}\right) \quad \text{S27}$$

The timing one would infer under the assumption of constant mutation rate is therefore:

$$\tilde{t}_{k,i}^{est} = \frac{\tilde{t}_{k,i}^{true} - (1-\rho)\tilde{t}_{k,1}^{true}}{1 - (1-\rho)\tilde{t}_{k,1}^{true}} \quad \text{S28}$$

Rearranging equation S28 gives the true chronological timing of any CNA event $i$ after the first:

$$\tilde{t}_{k,i}^{true} = \tilde{t}_{k,i}^{est}\left(1 - (1-\rho)\tilde{t}_{k,1}^{true}\right) + (1-\rho)\tilde{t}_{k,1}^{true} \quad \text{S29}$$

$$\tilde{t}_{k,i}^{true} = \tilde{t}_{k,i}^{est} + \left(1 - \tilde{t}_{k,i}^{est}\right)(1-\rho)\tilde{t}_{k,1}^{true} \quad \text{S30}$$

Finally, substituting our expression for $\tilde{t}_{k,1}^{true}$ in S25 into S30 yields a solution for the true chronological timing of any CNA event purely in terms of the estimated constant rate timings and $\rho$:

$$\tilde{t}_{k,i}^{true} = \frac{\rho\tilde{t}_{k,i}^{est} + (1-\rho)\tilde{t}_{k,1}^{est}}{\rho + (1-\rho)\tilde{t}_{k,1}^{est}} \quad \text{S31}$$

Validation of analytic correction

The full CARBINE model contains additional complexity compared to the simplified model version used to derive the correction factor given in S31 (e.g. the time since the MRCA and the term corresponding to the number of diploid mutations observed), but a proper correction for the full model would be more difficult to derive analytically. Hence, we sought to validate the accuracy of this approximate correction when applied to the full CARBINE model using simulated data. We used a modified form of our simulator incorporating an additional free parameter, $\rho$, as above. Setting $\rho = \frac{1}{3}$ (i.e. a 3x acceleration following the first CNA), we ran CARBINE on our generated simulations and indeed observed a systematic underestimate of the true CNA time due to CARBINE's constant mutation rate assumption (supplementary fig. 4A). However, applying the analytic correction factor with $\rho = \frac{1}{3}$ allowed us to faithfully recover a non-biased estimate of the CNA time (supplementary fig. 4B). Hence, the analytic correction factor is applicable despite the additional model complexity within CABRINE.
